# Rise of escitalopram and the fall of citalopram

**DOI:** 10.1101/2023.05.07.23289632

**Authors:** Luke R. Cavanah, Parita Ray, Jessica L. Goldhirsh, Leighton Y. Huey, Brian J. Piper

## Abstract

**Introduction:** Citalopram and escitalopram are among the most used medications and are key treatments for many psychiatric disorders. Previous findings suggest citalopram and escitalopram prescription rates are changing because of the patent for citalopram ending as opposed to evidence of a clear therapeutic advantage, which is called evergreening. This retrospective study focuses on characterizing the chronologic and geographic variation in the use of citalopram and escitalopram among US Medicaid and Medicare patients. We hypothesized that prescription rates of citalopram will decrease with a concurrent increase in escitalopram, consistent with evergreening.

**Methods:** Citalopram and escitalopram prescription rates and costs per state were obtained from the Medicaid State Drug Utilization Database and Medicare Provider Utilization and Payment Data. Annual prescription rates outside a 95% confidence interval were considered significantly different from the average.

**Results:** Overall, a decreasing trend for citalopram and an increasing trend for escitalopram prescription rates were noted in both Medicare and Medicaid patients. Cost differences between generic and brand were noted for both drugs, with generic forms being cheaper compared to the brand-name version.

**Discussion:** Despite limited evidence suggesting that citalopram and escitalopram have any meaningful differences in therapeutic or adverse effects, there exists a noticeable decline in the use of citalopram that cooccurred with an increase in escitalopram prescribing, consistent with our hypothesis. Moreover, among these general pharmacoepidemiologic trends exists significant geographic variability. There was disproportionate spending (relative to their use) on the brand versions of these medicines compared to their generic forms.

## Introduction

Consistently, citalopram and escitalopram are two of the fifty most commonly prescribed medications in the US (Kane, 2021a, 2021b, 2021c, 2021d, 2021e). Citalopram and escitalopram were FDA-approved for the treatment of major depressive disorder (MDD), and escitalopram is additionally FDA-approved for the treatment of generalized anxiety disorder (GAD). Both medications are used off-label for numerous other psychiatric conditions, such as obsessive-compulsive disorder, panic disorder, post-traumatic stress disorder, premenstrual dysphoric disorder, and social anxiety disorder. Citalopram, which is available in generic form and as Celexa ®, was introduced in 1998, and it is a racemic mixture of the S(+)-enantiomer (escitalopram) and the R(−)-enantiomer (R-citalopram) (Lochmann & Richardson, 2019; Milne & Goa, 1991). Although both citalopram and escitalopram are members of the “selective” reuptake inhibitors class, these agents showed significant, and equivalent, affinity of the sigma_1_ receptors (Sánchez et al., 2004). Citalopram’s affinity for the histamine (H_1_) receptor (257 ± 2.8 nm) was described as “weak” which is only slightly greater than that of escitalopram’s (1,500 ± 780 nm) (Sánchez et al., 2004; Stahl, 2020). Some studies suggest that the S-enantiomer, but not the R-enantiomer, was responsible for the therapeutic effect of citalopram (Hogg & Sánchez, 1999; Hyttel et al., 1992). Escitalopram and citalopram generally produced equivalent effects on six animal models of depression, anxiety, and aggression although citalopram was slightly more potent (Sanchez et al. 2004). Moreover, there is some evidence that perhaps the R-enantiomer results in a slightly diminished therapeutic effect of the S-enantiomer, leading to the introduction of escitalopram in 2002 (Sánchez et al., 2004). Escitalopram is now available in generic form and as Lexapro ®. The general conclusion from both basic and clinical research was that the differences between citalopram and escitalopram were of potency and not efficacy.

Part of the reason citalopram and escitalopram are so commonly used is due to their relatively high tolerability. Both patients taking citalopram and escitalopram commonly report adverse effects of dry mouth, headache, nausea and vomiting, diarrhea, insomnia, and ejaculatory problems (Landy et al., 2022; Sharbaf Shoar et al., 2022; Waugh & Goa, 2003). Rarely, these medicines may result in serious conditions, such as syndrome of inappropriate antidiuretic hormone or serotonin syndrome (Landy et al., 2022; Sharbaf Shoar et al., 2022). Interestingly, patients taking citalopram may have adverse effects of QTc prolongation at a rate, albeit small, that was mildly greater than patient who take escitalopram (Funk & Bostwick, 2013).

Citalopram and escitalopram are frequently used among Medicaid and Medicare patients, emphasizing the importance of investigation of these medications within these populations. Notably, psychopharmaceuticals comprised the second most prescribed medication type for outpatient Medicaid patients (Dolan, 2021). Psychological disorders result in a huge economic burden that is estimated to continue to increase (Doran & Kinchin, 2019). Furthermore, mental disorders are one of the most commonly treated disorders within the top five percent of spenders (Mitchell, 2022). Recent research has begun to elucidate how the COVID-19 pandemic has contributed to increased frequency of these disorders (Ettman et al., 2020; Lakhan et al., 2020; Salari et al., 2020; Talevi et al., 2020), again underscoring the need to understand the pharmacoepidemiology of treatments, especially those as ubiquitous as citalopram and escitalopram.

“Evergreening” refers to the deceptive techniques, such as incremental drug modification, that pharmaceutical companies use to continue their monopoly over a drug’s rights (Bansal et al., 2009; Dwivedi et al., 2010; Hemphill & Sampat, 2012). There is limited evidence that supports that citalopram is a more effective or tolerable alternative to escitalopram, yet an earlier meta-analysis suggests that citalopram/escitalopram prescription rates follow the patterns that would be expected for the case of evergreening (Alkhafaji et al., 2012).

The purpose of this study was to examine patterns in citalopram and escitalopram prescription rates throughout the US among Medicaid and Medicare patients. We hypothesized that citalopram prescription rates will decrease across the time interval with a concurrent increase in escitalopram prescription rates for both Medicaid and Medicare populations. This finding would suggest that evergreening has continued to be observed with these medications. A secondary objective was to examine state-level disparities in use of these antidepressants. The high frequency of use of these medicines makes these findings particularly informative.

## Methods

### Procedures

Citalopram and escitalopram prescription rates and costs were obtained for Medicaid and Medicare. Medicaid and Medicare data were assessed quarterly and annually (2015-2020), respectively, due to the information publicly available. We evaluated the Medicaid State Drug Utilization database (Medicaid, n.d.) and Medicare Provider Utilization and Payment Data (Centers for Medicare & Medicaid Services, n.d.) for citalopram and escitalopram prescription rates per state. We evaluated the Medicaid National Drug Utilization database for citalopram and escitalopram national prescription rates per quarter. Prescription rates were reported per thousand enrollees. Procedures were defined as exempt by the Geisinger IRB.

### Data Analysis

Patterns in the number of national prescriptions of generic, brand, and their sum were compared for both citalopram and escitalopram. One-sample z-tests were conducted to determine if any states’ annual prescription rates of either of these medications were significantly different from the state average for that respective year for the respective program. The ratio of total Medicaid spending and Medicare spending for generic versus brand of these medications was also calculated. Average costs per prescription of citalopram and escitalopram were also assessed. We analyzed the data using Excel and constructed figures using GraphPad Prism and Heatmapper (Babicki et al., 2016).

## Results

### Medicaid

Figure 1 shows -26.0% decreasing prescriptions for citalopram from 2015 to 2019 and +41.2% increasing rate of prescriptions for escitalopram. The brand names for both citalopram (−42.3%) and escitalopram (−85.9%) had a decreasing rate of prescriptions from 2015 to 2020. Analysis of generic and brand name prescriptions combined revealed similar rates of prescriptions as their respective generic version amongst Medicaid patients.

**Figure 1.**
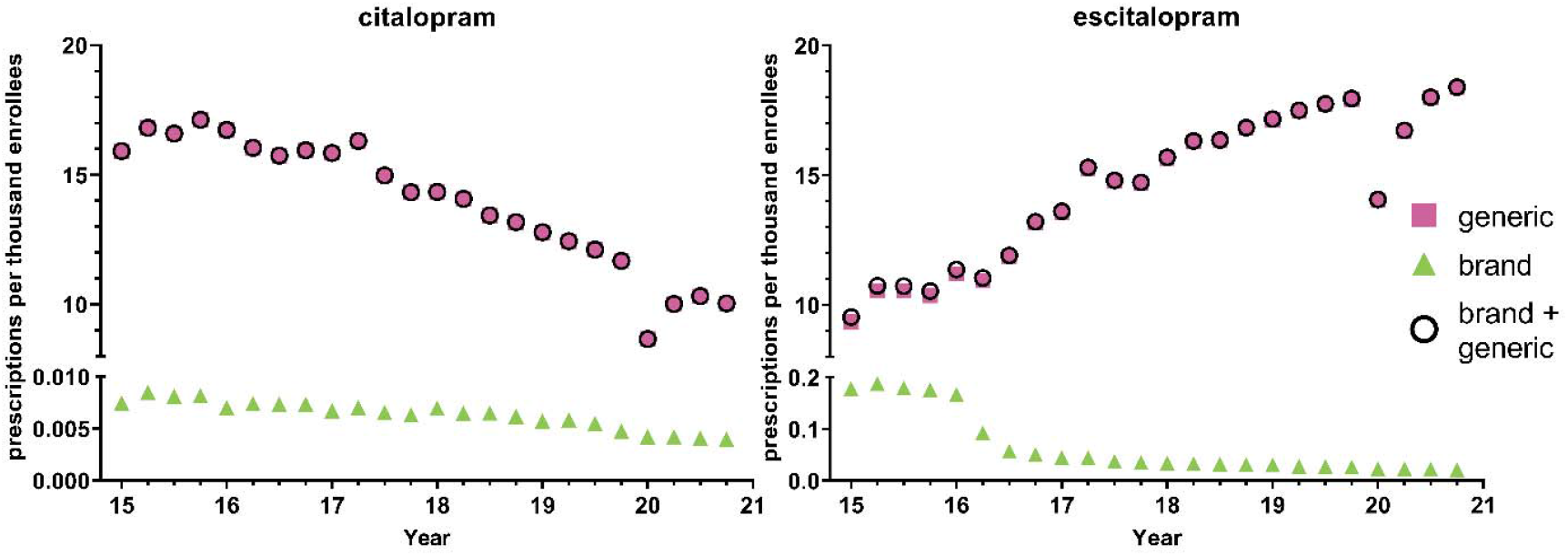
National Medicaid population-corrected prescriptions for citalopram and escitalopram 2015-2020.

Quarterly examination revealed a decrease in prescription rates in the first quarter of 2020 relative to the fourth quarter of 2019, a -25.9% reduction for citalopram and -21.7% reduction for escitalopram, followed by an increase in prescription rates for both citalopram and escitalopram during the later quarters. Prescriptions for the brand names of the respective drugs also showed a gradual decrease throughout the years, except for Lexapro, which displayed a - 55.6% decrease in the first quarter of 2016 (Figure 1). Overall, the prescribing rates for the brand name of the respective drugs were significantly lowered compared to its generic counterpart (Figure 1). Analysis of cost per prescription data shows an average of a 50-fold increase in the price for the brand names of citalopram and escitalopram compared to its generic counterparts.

Table 1 shows pronounced disparities by state. On average, there was a nineteen-fold difference between the highest and lowest states for escitalopram relative to an eleven-fold difference for citalopram. Figure 2 and Supplemental Figures 1-5 display state-level variations in the prescription for citalopram amongst Medicaid patients. In 2020, Kentucky (117.2), the highest prescribing state, was 9.9-fold greater than Arizona (11.8), the lowest prescribing state (Figure 2). Kentucky (117.2) and West Virginia (106.4) showed statistically significant (p<0.05) higher rates of prescriptions compared to the national average (Figure 2). Kentucky and West Virginia continued to be statistically significantly higher in the rates of prescriptions compared to the national average for 2016, 2017, and 2018 (Supplemental Figures 2-4). For 2015 and 2019, only West Virginia (155.8) was statistically significantly higher compared to the national average (Supplemental Figure 5).

**Table 1.** Pronounced state level disparities in Medicaid and Medicare prescriptions, per 1,000 enrollees, for citalopram and escitalopram. Abbreviations for the highest and lowest states are in superscript (AR: Arkansas, AZ: Arizona, CT: Connecticut, DC: District of Columbia, KY: Kentucky, NM: New Mexico, RI: Rhode Island, SC: South Carolina, WV: West Virginia).

|  | citalopram |  |  | escitalopram |  |  |
| --- | --- | --- | --- | --- | --- | --- |
| <u>Medicaid</u> | <u>Highest</u> | <u>Lowest</u> | <u>Ratio</u> | <u>Highest</u> | <u>Lowest</u> | <u>Ratio</u> |
| 2015 | 167.3<sup>WV</sup> | 7.3<sup>RI</sup> | 22.9 | 122.7<sup>CT</sup> | 1.5<sup>RI</sup> | 81.8 |
| 2016 | 160.6<sup>WV</sup> | 22.3<sup>HI</sup> | 7.2 | 128.5<sup>WV</sup> | 19.2<sup>SC</sup> | 6.7 |
| 2017 | 178.6<sup>KY</sup> | 23.6<sup>DC</sup> | 7.6 | 132.7<sup>WV</sup> | 24.8<sup>DC</sup> | 5.4 |
| 2018 | 178.5<sup>KY</sup> | 20.4<sup>DC</sup> | 8.8 | 145.4<sup>WV</sup> | 24.5<sup>DC</sup> | 5.9 |
| 2019 | 152.4<sup>KY</sup> | 18.3<sup>SC</sup> | 8.3 | 155.8<sup>WV</sup> | 26.7<sup>NM</sup> | 5.8 |
| 2020 | 117.2<sup>KY</sup> | 11.8<sup>AZ</sup> | 9.9 | 144.4<sup>WV</sup> | 27.8<sup>DC</sup> | 5.2 |
| Mean | 163.5 | 17.3 | 10.8 | 138.3 | 20.8 | 18.5 |
| <u>Medicare</u> |  |  |  |  |  |  |
| 2015 | 453.4<sup>AR</sup> | 100.2<sup>HI</sup> | 4.5 | 361.4<sup>CT</sup> | 109.0<sup>HI</sup> | 3.3 |
| 2016 | 436.2<sup>AR</sup> | 92.1<sup>HI</sup> | 4.7 | 367.7<sup>CT</sup> | 107.7<sup>HI</sup> | 3.4 |
| 2017 | 403.2<sup>AR</sup> | 80.2<sup>HI</sup> | 5.0 | 365.5<sup>CT</sup> | 103.7<sup>HI</sup> | 3.5 |
| 2018 | 369.6<sup>AR</sup> | 75.0<sup>HI</sup> | 4.9 | 361.6<sup>CT</sup> | 99.8<sup>HI</sup> | 3.6 |
| 2019 | 328.4<sup>AR</sup> | 64.1<sup>HI</sup> | 5.1 | 351.4<sup>CT</sup> | 95.2<sup>HI</sup> | 3.7 |
| 2020 | 295.1<sup>AR</sup> | 59.9<sup>HI</sup> | 4.9 | 341.5<sup>CT</sup> | 96.4<sup>HI</sup> | 3.5 |
| Mean | 381.0 | 78.6 | 4.9 | 358.2 | 102.0 | 3.5 |

**Figure 2.**
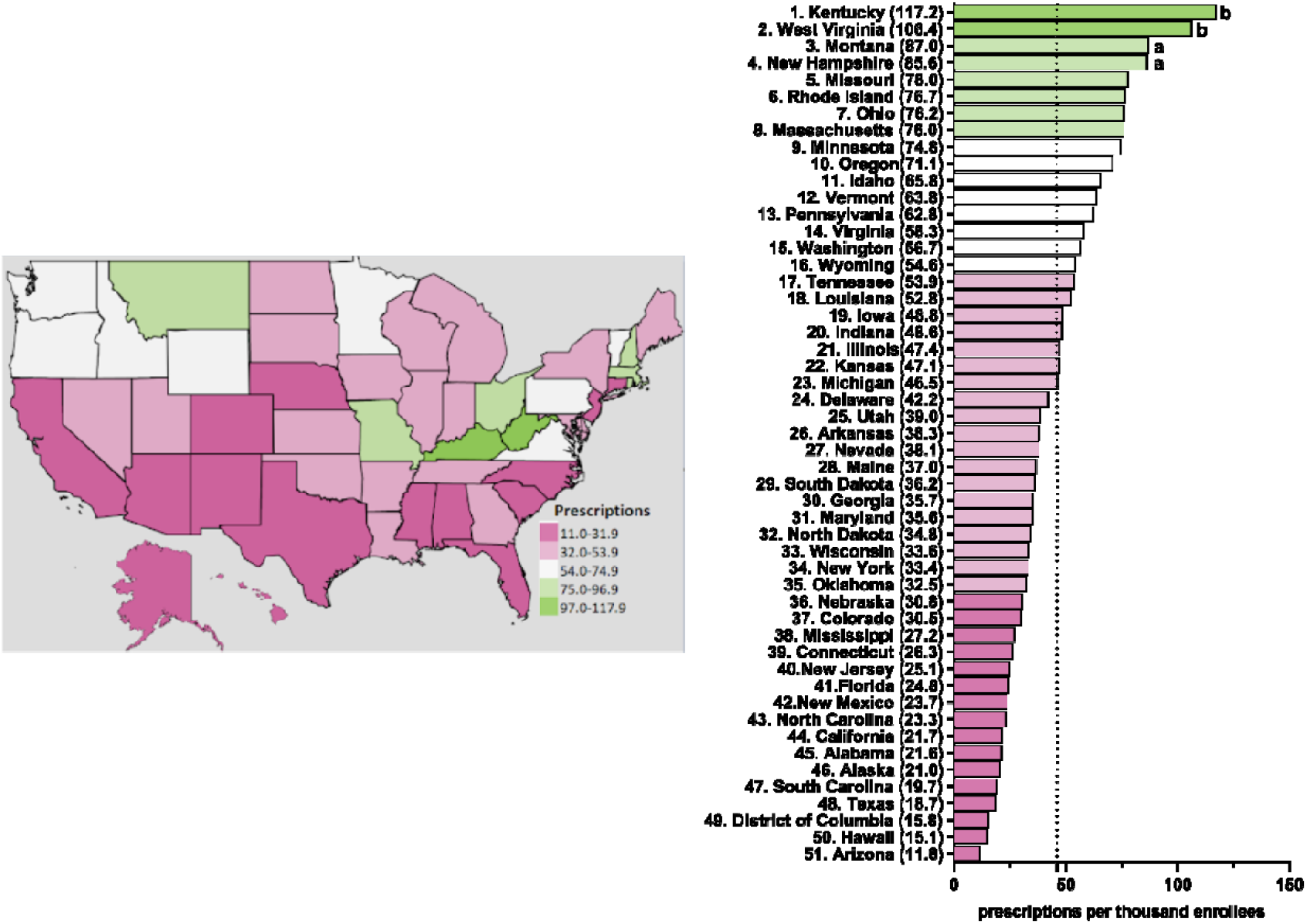
Citalopram prescriptions per thousand Medicaid enrollees heatmap (left) and population-corrected prescription rate per state (right) in 2020. ^a^ indicates ≥1.50 SD (24.0) from the mean (46.1). ^b^ indicates ≥1.96 SD from the mean.

Figure 3 and Supplemental Figures 6-10 display state-level variations in the prescription for escitalopram amongst Medicaid patients. In 2020, West Virginia (144.4), the highest prescribing state, was 5.2-fold greater compared to the lowest prescribing area, the District of Columbia (27.8) (Figure 3). West Virginia was consistently among the highest statistically significant prescribing states of escitalopram from 2015-2020 (Figure 3 and Supplemental Figures 6-10). In 2018, North Dakota was also among the top prescribers of escitalopram (Supplemental Figure 9). Additionally, in 2015 and 2016, Connecticut and Iowa were among the highest statistically significant prescribing states compared to the national average (Supplemental Figures 6-7).

**Figure 3.**
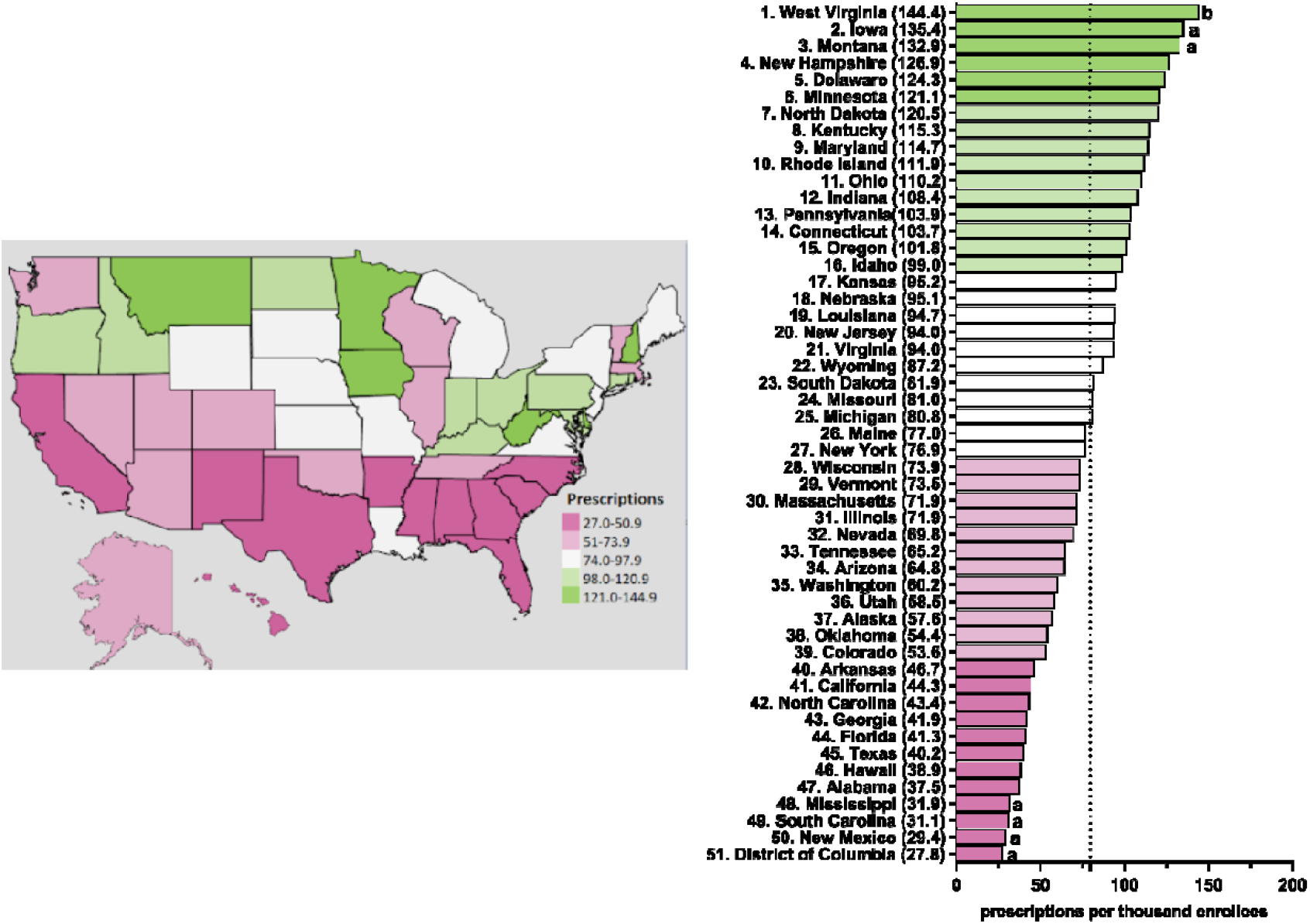
Escitalopram prescriptions per thousand Medicaid enrollees heatmap (left) and population-corrected prescription rate per state (right) in 2020. ^a^ indicates ≥1.50 SD (31.6) from the mean (79.6). ^b^ indicates ≥1.96 SD from the mean.

### Medicare

Figure 4 displays a –24.5% decreasing rate of prescriptions for citalopram from 2015 to 2020 and +5.61% increasing rate of prescriptions for escitalopram for Medicare patients. The brand name versions of these drugs showed a gradual decrease in prescription rates. Like Medicaid patients, analysis of generic and brand name prescriptions combined revealed similar rates of prescriptions as their respective generic versions in Medicare recipients.

**Figure 4.**
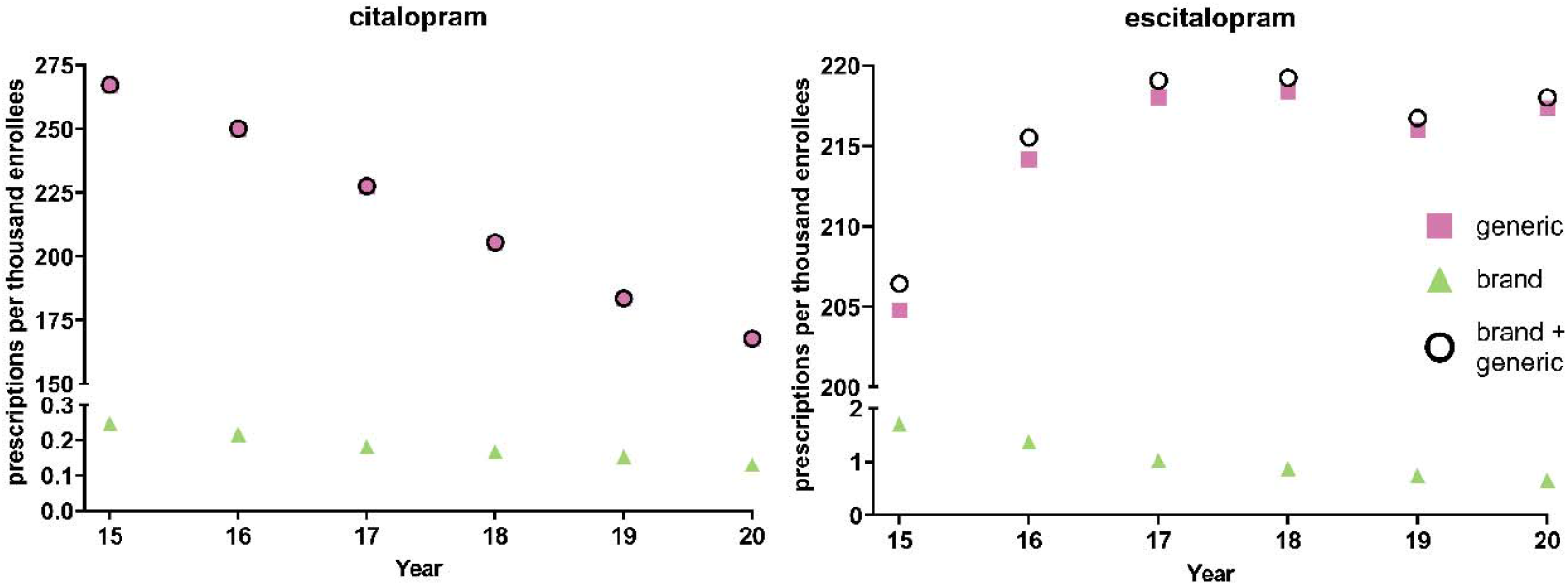
National Medicare population-corrected prescriptions for citalopram and escitalopram 2015-2020.

Table 1 shows appreciable (three to four-fold) state-level differences between the highest (Arkansas and Connecticut) and lowest (Hawaii) states. Figure 5 and Supplemental Figures 11-15 depict state-level variations in the prescription for citalopram amongst Medicare enrollees. From 2015-2020, Arkansas was the leading prescribing state of citalopram, with it being statistically significantly higher in the rates of prescriptions compared to the national average. Conversely, Hawaii has statistically been the lowest prescribing state compared to the national average from 2015-2020 (Figure 5 and Supplemental Figures 11-15). In addition, New Jersey was statistically the lowest prescribing state of citalopram compared to the national average from 2015-2018 and 2020 (Figure 5 and Supplemental Figures 11-14).

**Figure 5.**
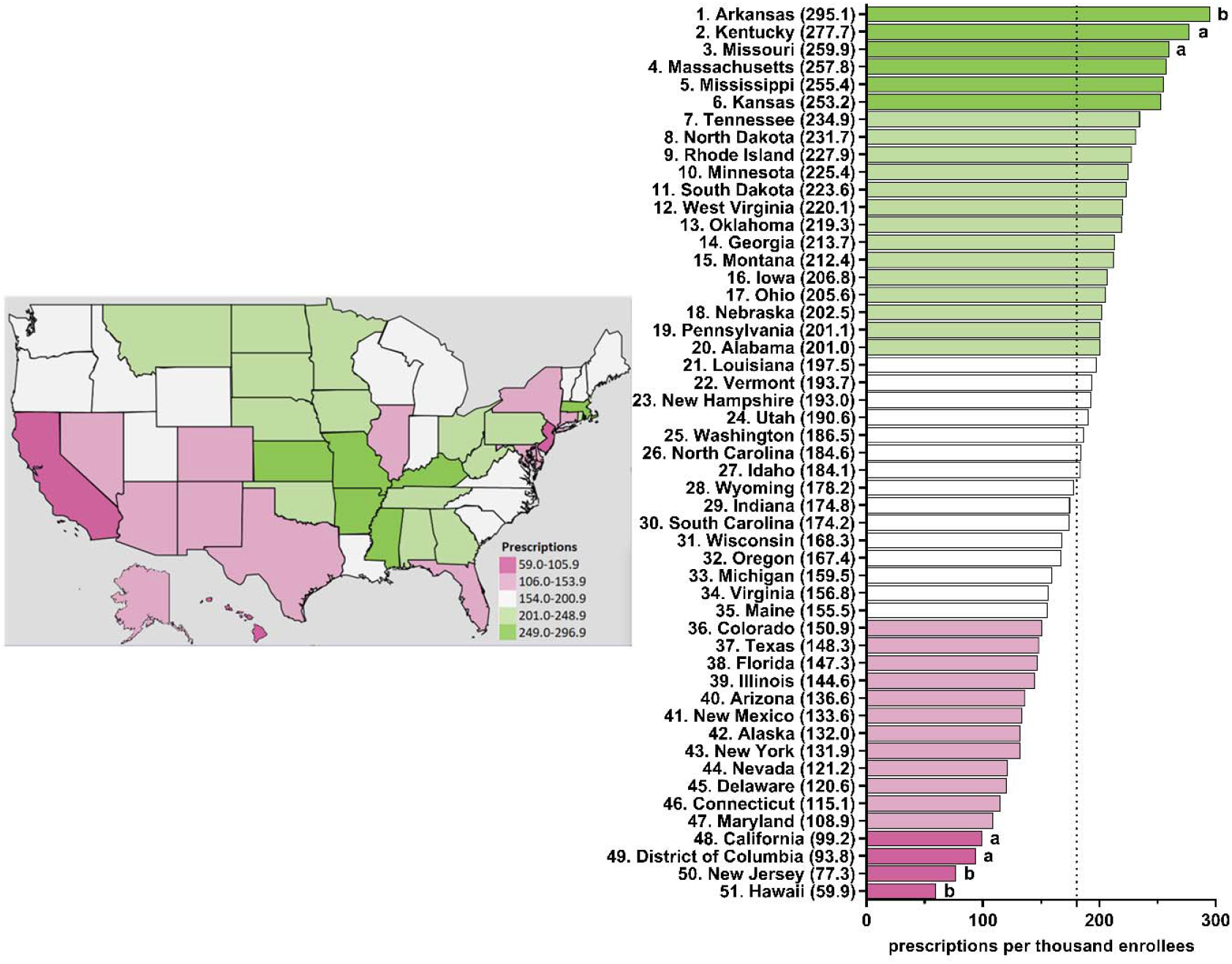
Citalopram prescriptions per thousand Medicare enrollees heatmap (left) and population-corrected prescription rate per state (right) in 2020. ^a^ indicates ≥1.50 SD (51.8) from the mean (180.6). ^b^ indicates ≥1.96 SD from the mean.

Figure 6 and Supplemental Figures 16-20 display state-level variations in the prescription for escitalopram amongst Medicare enrollees. From 2015-2020, Connecticut has consistently been the top prescriber of escitalopram and statistically higher compared to the national average (Figure 6 and Supplemental Figures 16-20). Additionally, North Dakota was statistically higher compared to the national average in 2019 and 2020 (Figure 6 and Supplemental Figure 20). Conversely, Hawaii was statistically the lowest prescriber of escitalopram compared to the national average from 2016-2020 (Figure 6 and Supplemental Figures 17-20).

**Figure 6.**
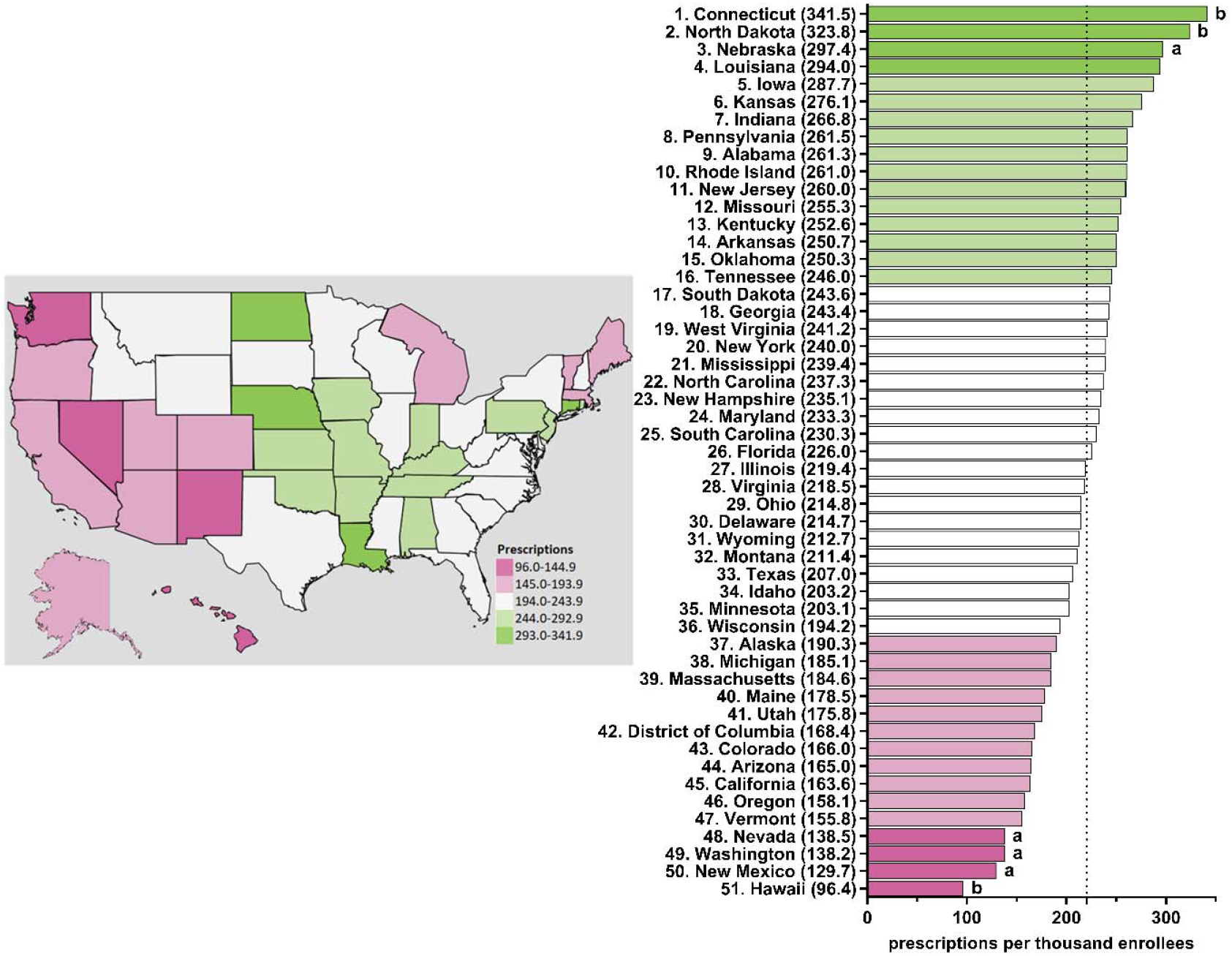
Escitalopram prescriptions per thousand Medicare enrollees heatmap (left) and population-corrected prescription rate per state (right) in 2020. ^a^ indicates ≥1.50 SD (49.9) from the mean (220.6). ^b^ indicates ≥1.96 SD from the mean.

## Discussion

Between 2015-2020, the US demonstrated an increase in prescriptions of escitalopram and a corresponding decrease in prescriptions of citalopram amongst both Medicare and Medicaid patients. Despite both citalopram and escitalopram having equal efficacy (Sánchez et al., 2004) and similar adverse effects (Landy et al., 2022; Sharbaf Shoar et al., 2022; Waugh & Goa, 2003), we identified declining rates of prescription of citalopram. Analysis of the Medical Expenditure Panel Survey revealed 30.2 million prescriptions for citalopram and 17.0 million for escitalopram in 2013 (Kane, 2021a). This pattern had reversed to 18.5 million citalopram prescriptions and 30.6 million escitalopram prescriptions in 2020 (Kane, 2021e) which is congruent with the present findings. About one-fifth (19.7%) of the US gross domestic product in 2020 was spent on health care (Statista Research Department, 2022) and this country was an outlier relative to other developed countries (Wager et al., 2022). Examining the patterns in rates of prescription of these commonly prescribed medications provides us with an insight into evergreening and how it affects the pharmacological industry and the pharmacoepidemiology of treatments.

Additionally, we found pronounced differences in prescriptions rates of citalopram and escitalopram at the state-level. Anxiety and depressive disorders and the most common psychiatric conditions (American Psychiatric Association, 2022). An assumption of this study was that the prevalence of anxiety and depression would show relatively modest difference based on state of residence (e.g., between Connecticut and adjacent Rhode Island). However, prescriptions for escitalopram in 2015 were eighty-fold more common in Connecticut relative to Rhode Island for Medicaid. Further, Kentucky was frequently among the highest, and statistically elevated, prescriber of citalopram and escitalopram to both Medicaid and Medicare enrollees, along with West Virginia, Connecticut, and Arkansas. These findings extend upon earlier findings which found five-fold state level differences among Medicare patients receiving paroxetine (Cavanah et al., 2023) six-fold for prescription stimulants (Vaddadi et al., 2021) and twenty-fold for meperidine (Harrison et al., 2022).

Escitalopram was initially introduced in 2002 as the S-enantiomer, while citalopram was introduced four years earlier in 1998 (Lochmann & Richardson, 2019; Milne & Goa, 1991; Sánchez et al., 2004). Clinical trials have determined that both escitalopram and citalopram significantly improve symptoms of depression and anxiety (Gorman et al., 2002) but that escitalopram may have a quicker onset and an overall greater effect on improving symptoms of anxiety and depression compared to citalopram among patients with major depressive disorder (Gorman et al., 2002). Although unlikely, this may be a possible explanation for the rise of escitalopram use over citalopram. The landmark Sequenced-Treatment-Alternatives to Relieve Depression (STAR*D) which found only a one-third remission rate with citalopram (Howland, 2008) may have tempered enthusiasm for the treatment of depression. While major drawback with SSRIs are the therapeutic lag and a black box warning for increased risk of suicidality among young-adults, further study will be needed to determine if novel agents like esketamine (Aguilar et al., 2023) will one day begin to supplant citalopram/escitalopram for depression.

Prescription rates of escitalopram in Medicaid enrollees almost doubled from 2015 to 2020 which may be explained by the concept of evergreening. Typically, the evergreened drug is released into the market before the patent expiration of the original drug. To compete with the original drug, one would expect the prices of the drugs to be competitive. We discovered that the cost of escitalopram per prescription was two-times higher compared to the cost of citalopram amongst Medicaid enrollees. For Medicare enrollees, the cost of escitalopram was five times higher than citalopram. The difference in costs with respect to the trends noted in the prescriptions of these medications suggests an alternate reason, which can be further assessed by another study.

In general, the use of SSRIs and total spending on them has increased from 1991 to 2018 (Alrasheed et al., 2021). The increased utilization of these drugs can also be explained by the rising prevalence of depression and the number of patients who seek pharmacological treatment. Generics were first introduced in 2001, which caused a shift in Medicaid to start the utilization of generic drugs over brand names (Alrasheed et al., 2021). This change allowed for a significant reduction in costs of these drugs and this pattern was noted in our study as well. It is also noteworthy that prescriptions of both citalopram and escitalopram to Medicaid patients showed transient, but appreciable (−21.7 – 25.9%) reductions during the initial period of the COVID-19 pandemic. Examining effects of COVID-19 pandemic on prescribing patterns of these medications and other psychotropics using other datasets is a meaningful future direction.

## Limitations

Some limitations to this report exist. First, although Medicaid and Medicare are large programs, these findings may not generalize to patients with private insurance (although see Kane, 2021). Second, further study with other data-sources will be necessary to determine if there are state-level differences in the access and utilization of evidence-based non-pharmacological treatments for anxiety (James et al., 2007) and depression (Li et al., 2018).

## Conclusion

In conclusion, citalopram used over the last five-years to US Medicaid and Medicare patients has decreased while escitalopram use continues to rise. The use of these SSRIs has also greatly shifted from brand to generic, which may be due to the high cost of brand-named drugs. Profound state-level variations in the prescriptions of these medications were noted in both Medicare and Medicaid patients. Future studies can explore the rising trends of these medications and explain the reason for the substantial state level differences among prescriptions.

## Data Availability

All data produced in the present study are available upon reasonable request to the authors

https://data.cms.gov/provider-summary-by-type-of-service/medicare-part-d-prescribers

## Acknowledgements

BJP was supported by HRSA (D34HP31025). Software used for this research was provided by NIEHS (T32-ES007060-31A1).

## Notes

**Disclosures:** BJP’s research is supported by the Pennsylvania Academic Clinical Research Center and the Health Resources and Services Administration (D34HP31025). Prior (2019 – 2021) osteoarthritis research was supported by Pfizer and Eli Lilly. The other authors have no disclosures.

### Competing Interest Statement

Disclosures: Research of BJP is supported by the Pennsylvania Academic Clinical Research Center and the Health Resources and Services Administration (D34HP31025). Prior (2019 to 2021) osteoarthritis research was supported by Pfizer and Eli Lilly. The other authors have no disclosures.

