## Supplemental Figures for "Rise of escitalopram and the fall of citalopram"

**Supplemental Figure 1.** Citalopram prescriptions per thousand Medicaid enrollees heatmap (left) and population-corrected prescription rate per state (right) in 2015. ^a^ indicates ≥1.50 SD (32.7) from the mean (71.2). ^b^ indicates ≥1.96 SD from the mean.

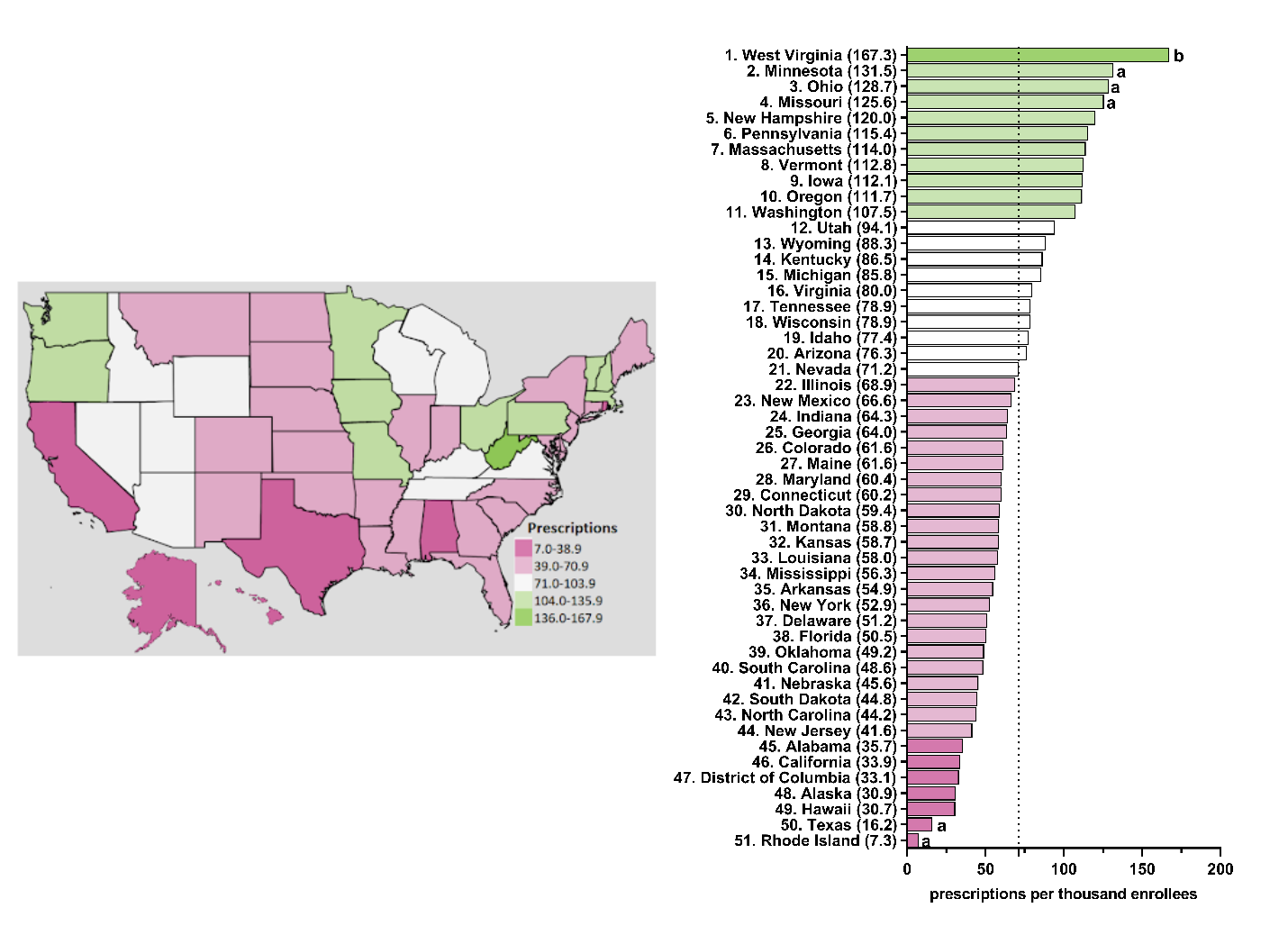

**Supplemental Figure 2.** Citalopram prescriptions per thousand Medicaid enrollees heatmap (left) and population-corrected prescription rate per state (right) in 2016. ^a^ indicates ≥1.50 SD (33.3) from the mean (71.6). ^b^ indicates ≥1.96 SD from the mean.

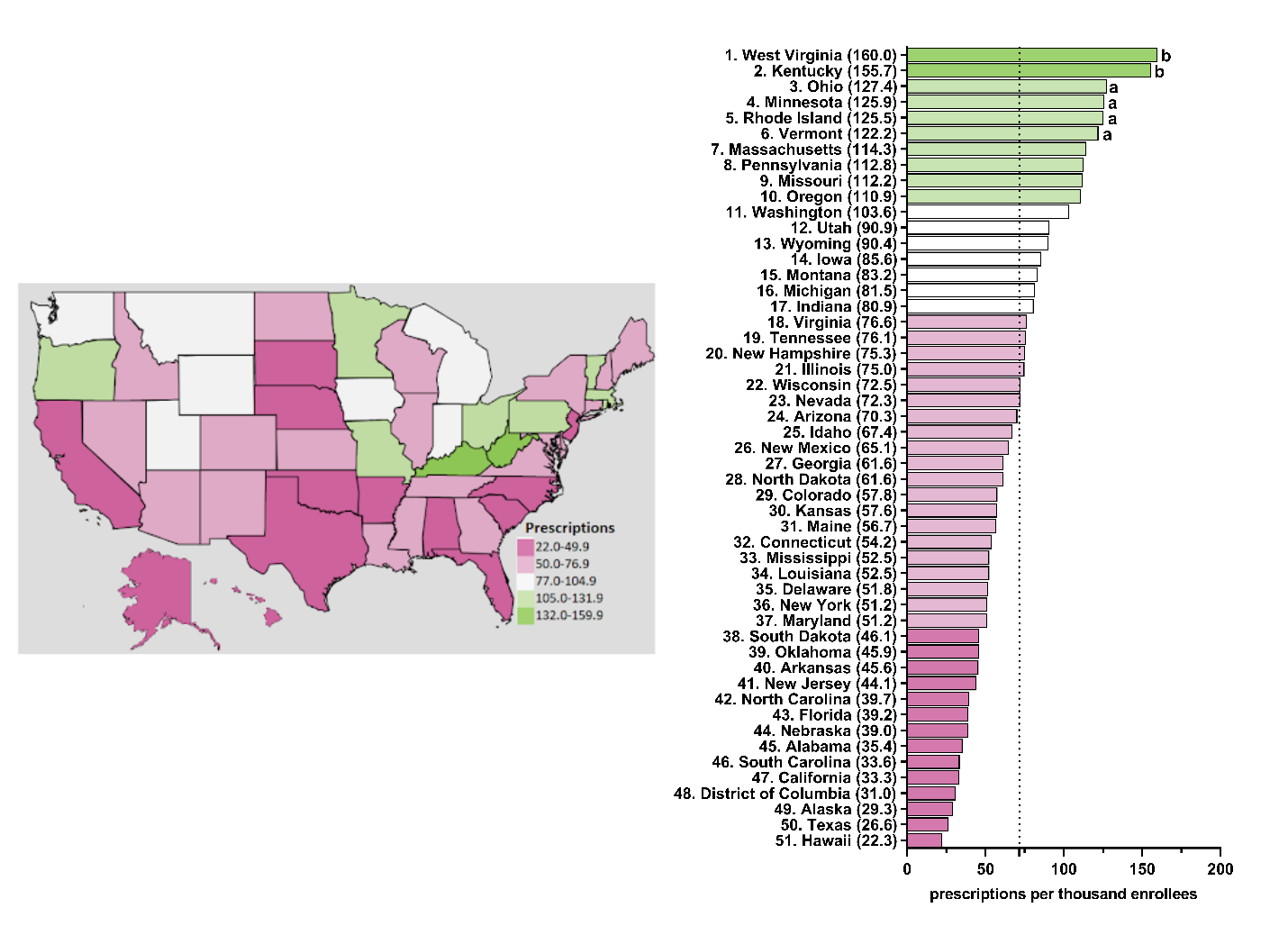

**Supplemental Figure 3.** Citalopram prescriptions per thousand Medicaid enrollees heatmap (left) and population-corrected prescription rate per state (right) in 2017. ^a^ indicates ≥1.50 SD (33.4) from the mean (67.4). ^b^ indicates ≥1.96 SD from the mean.

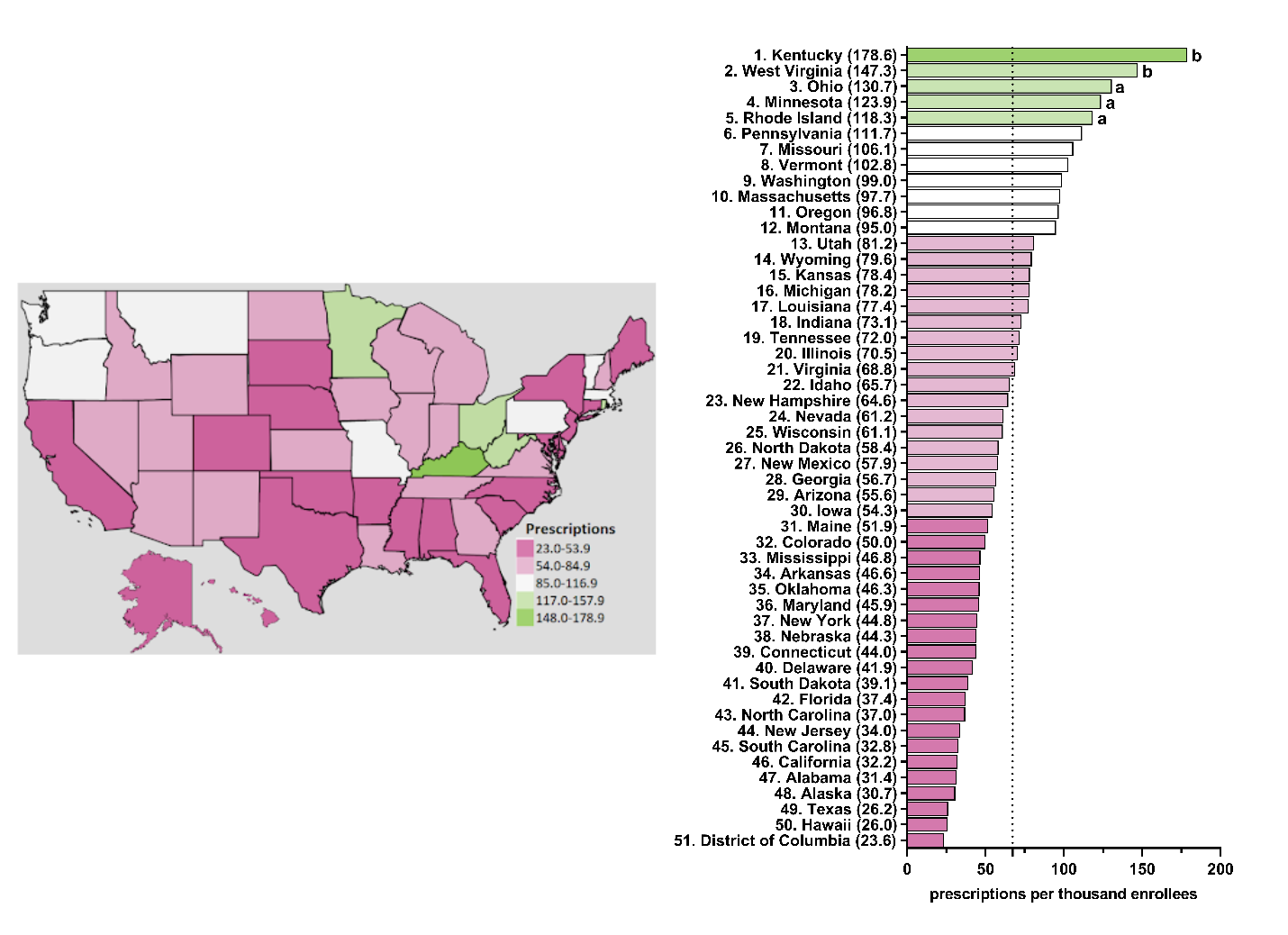

**Supplemental Figure 4.** Citalopram prescriptions per thousand Medicaid enrollees heatmap (left) and population-corrected prescription rate per state (right) in 2018. ^a^ indicates ≥1.50 SD (32.1) from the mean (61.0). ^b^ indicates ≥1.96 SD from the mean.

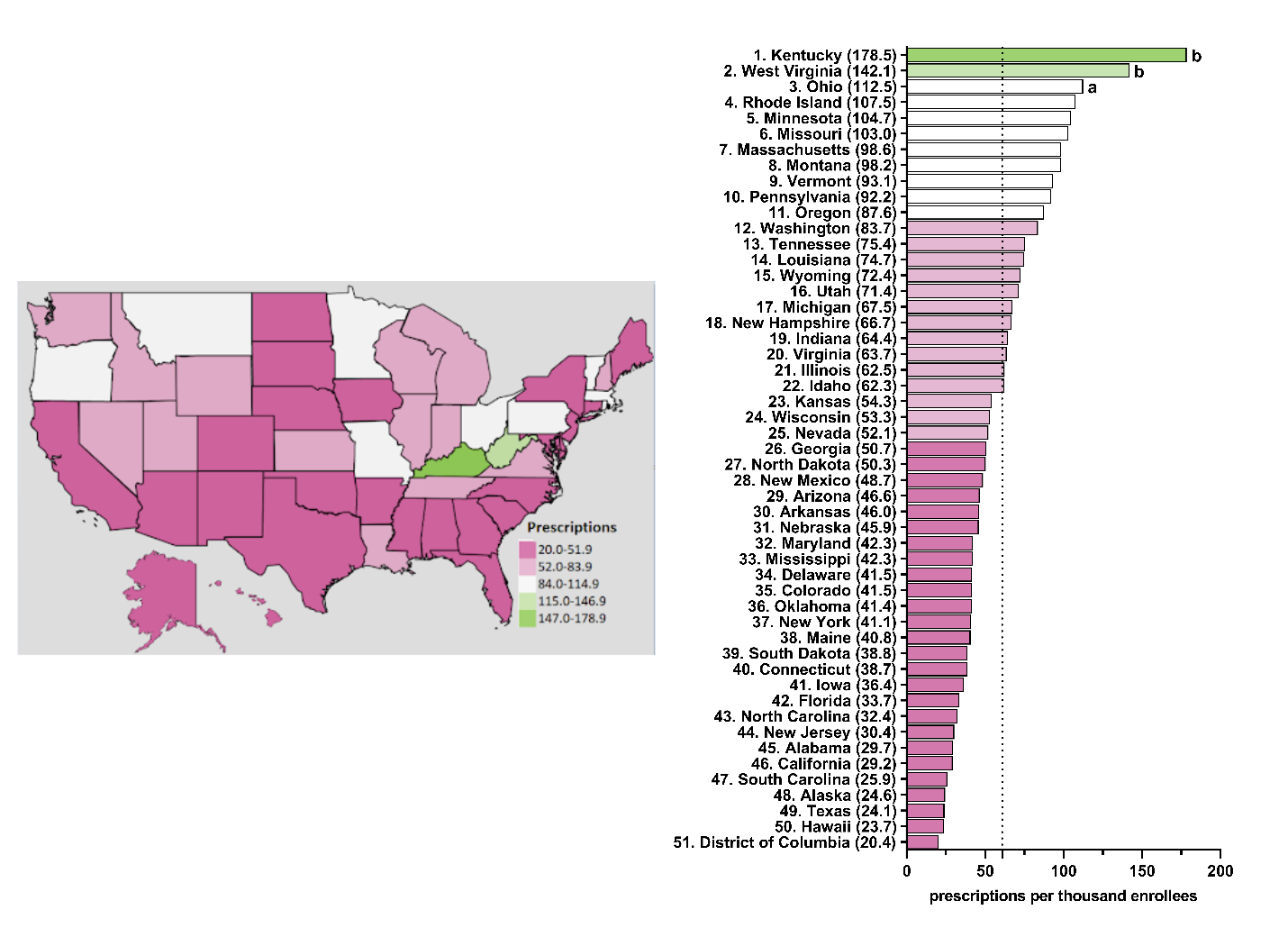

**Supplemental Figure 5.** Citalopram prescriptions per thousand Medicaid enrollees heatmap (left) and population-corrected prescription rate per state (right) in 2019. ^a^ indicates ≥1.50 SD (29.1) from the mean (55.4). ^b^ indicates ≥1.96 SD from the mean.

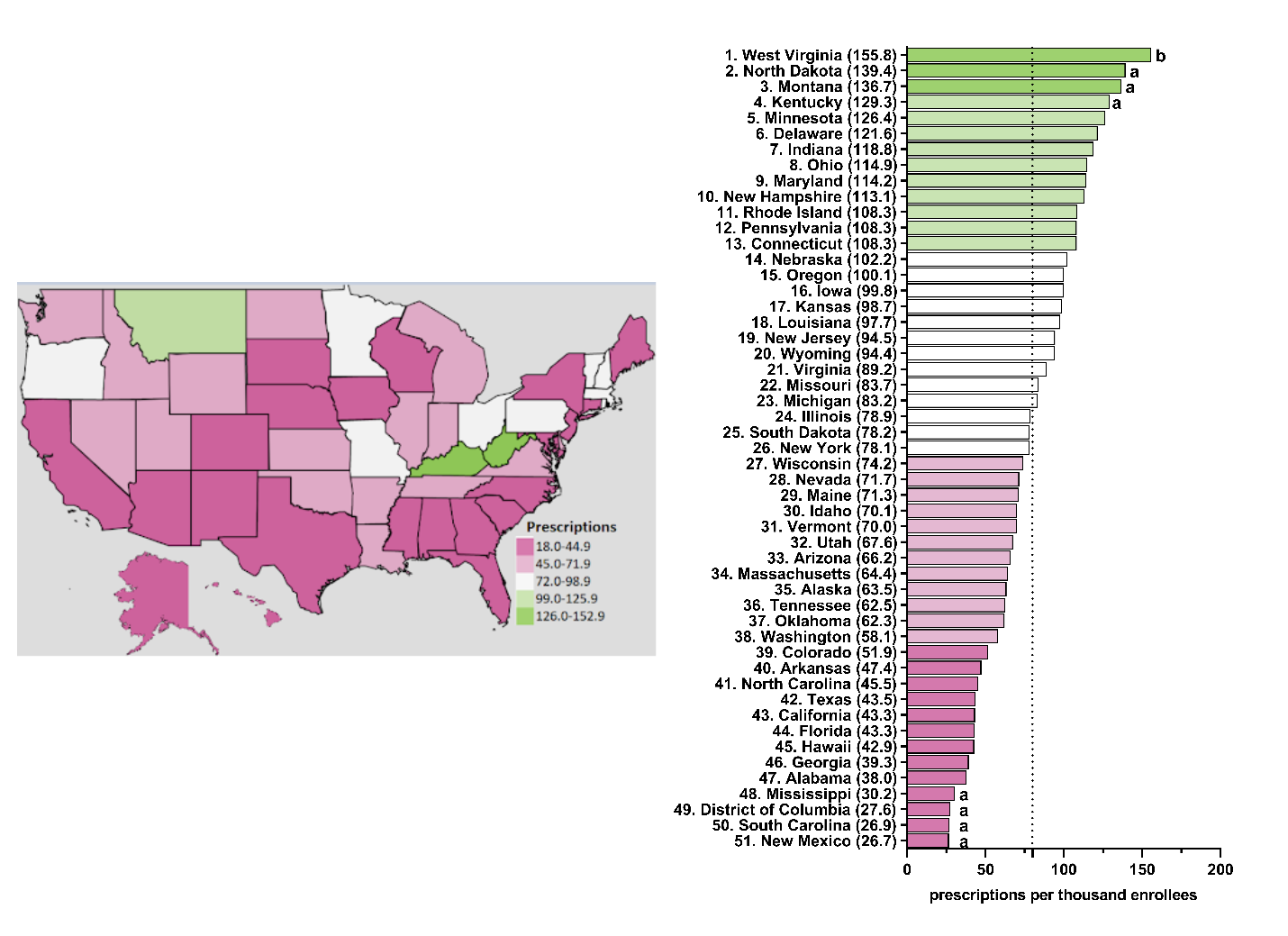

**Supplemental Figure 6.** Escitalopram prescriptions per thousand Medicaid enrollees heatmap (left) and population-corrected prescription rate per state (right) in 2015. ^a^ indicates ≥1.50 SD (26.2) from the mean (50.0). ^b^ indicates ≥1.96 SD from the mean.

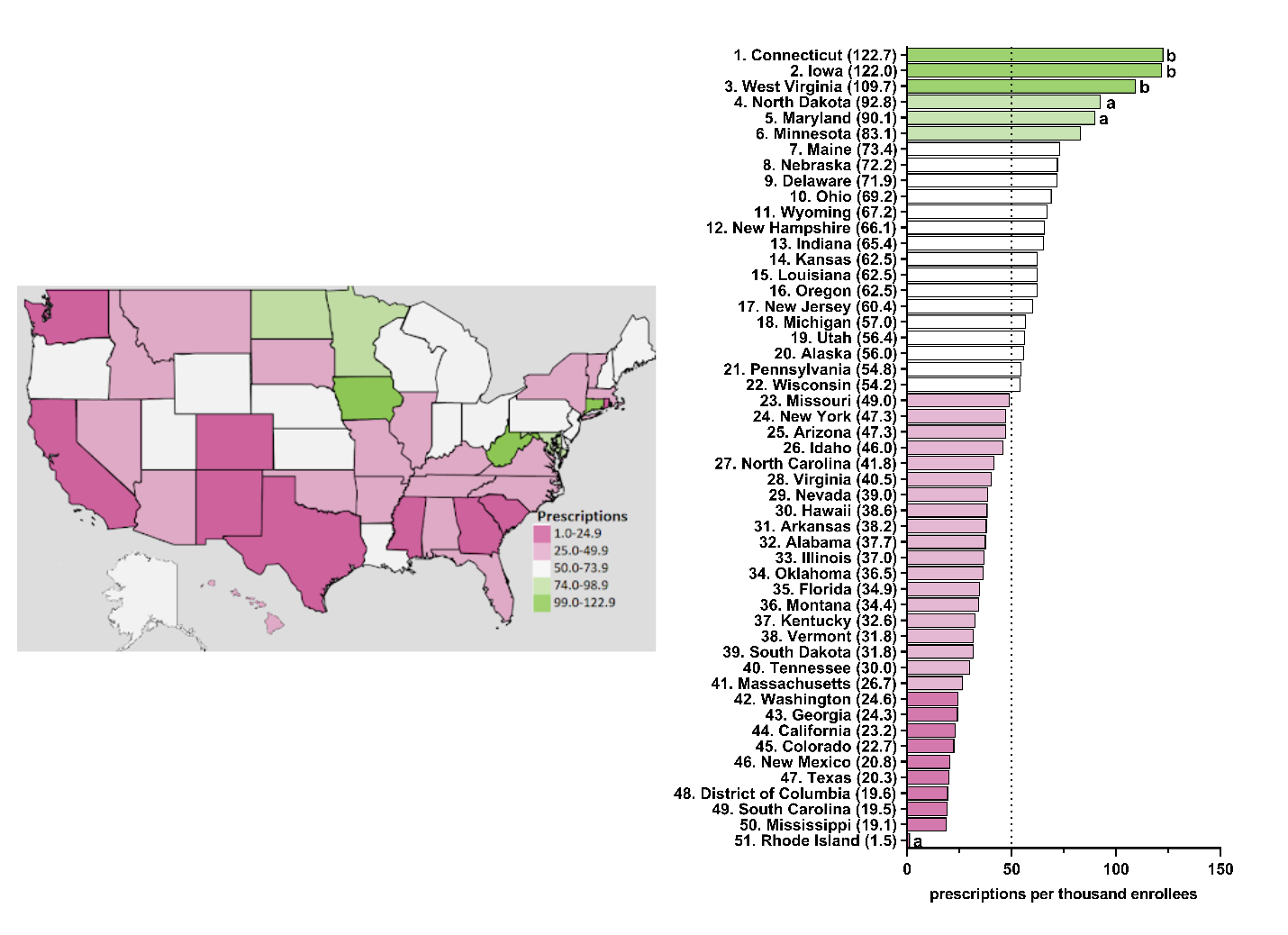

**Supplemental Figure 7.** Escitalopram prescriptions per thousand Medicaid enrollees heatmap (left) and population-corrected prescription rate per state (right) in 2016. ^a^ indicates ≥1.50 SD (27.2) from the mean (58.4). ^b^ indicates ≥1.96 SD from the mean.

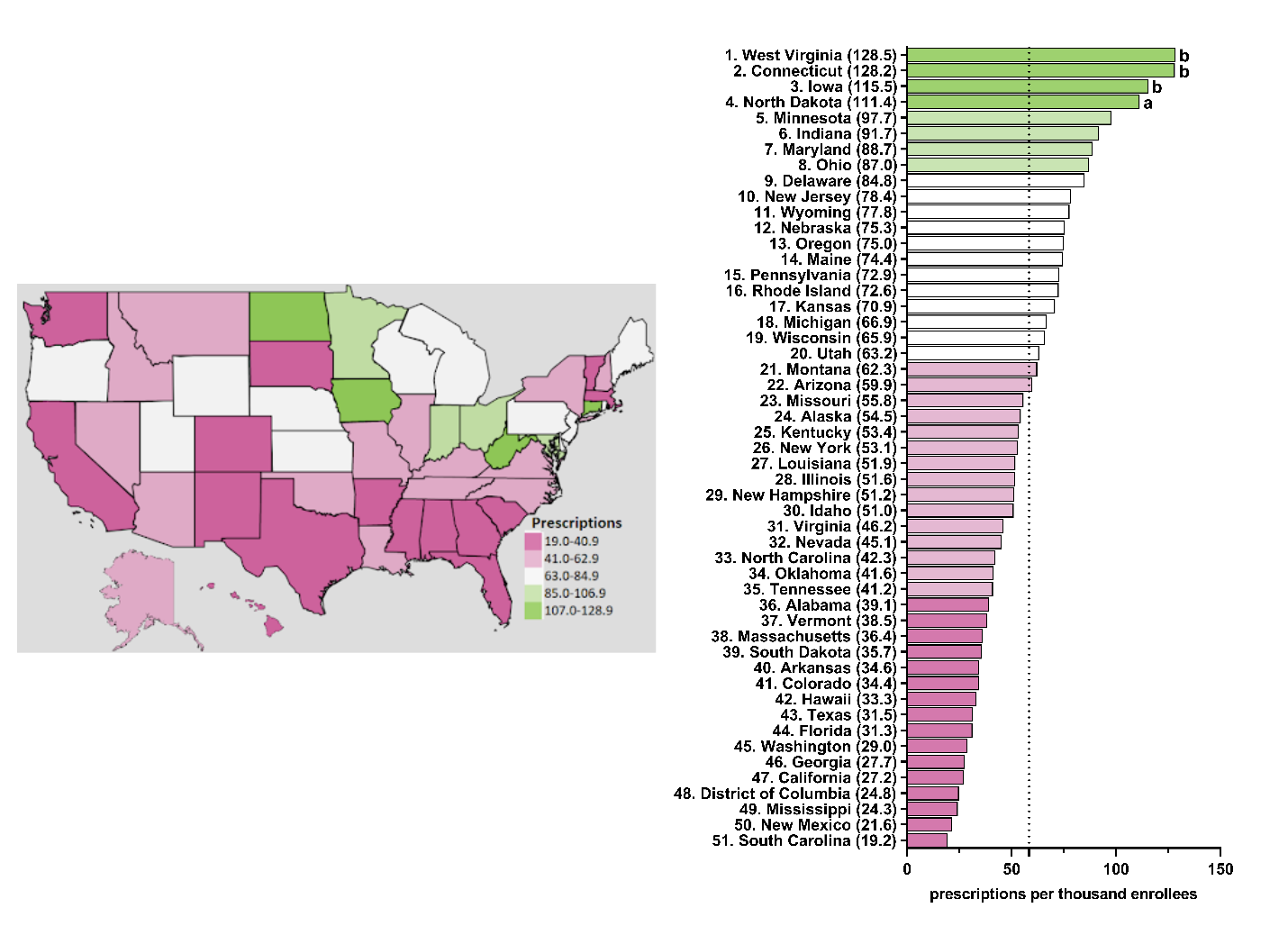

**Supplemental Figure 8.** Escitalopram prescriptions per thousand Medicaid enrollees heatmap (left) and population-corrected prescription rate per state (right) in 2017. ^a^ indicates ≥1.50 SD (27.6) from the mean (66.4). ^b^ indicates ≥1.96 SD from the mean.

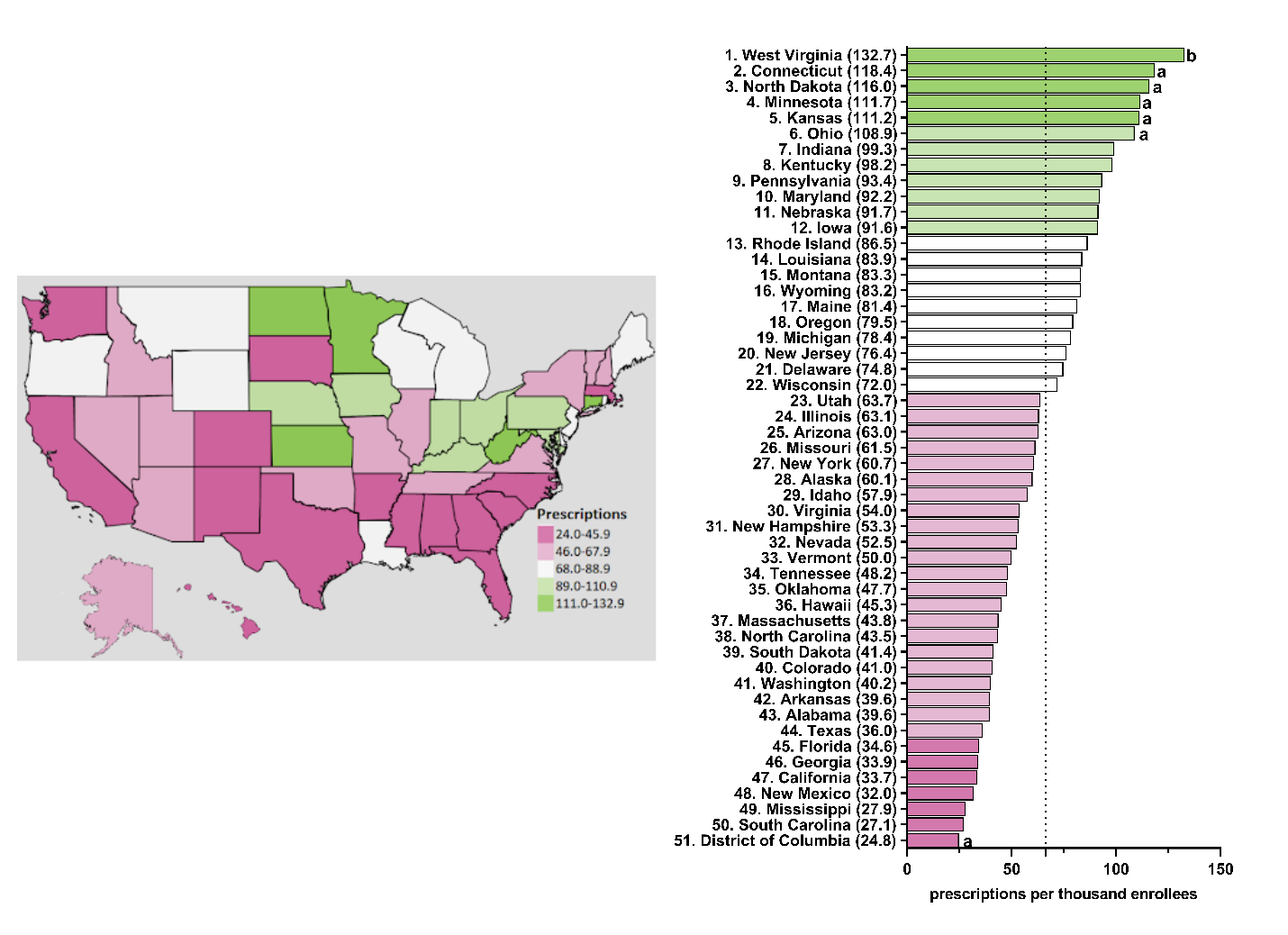

**Supplemental Figure 9.** Escitalopram prescriptions per thousand Medicaid enrollees heatmap (left) and population-corrected prescription rate per state (right) in 2018. ^a^ indicates ≥1.50 SD (28.8) from the mean (73.0). ^b^ indicates ≥1.96 SD from the mean.

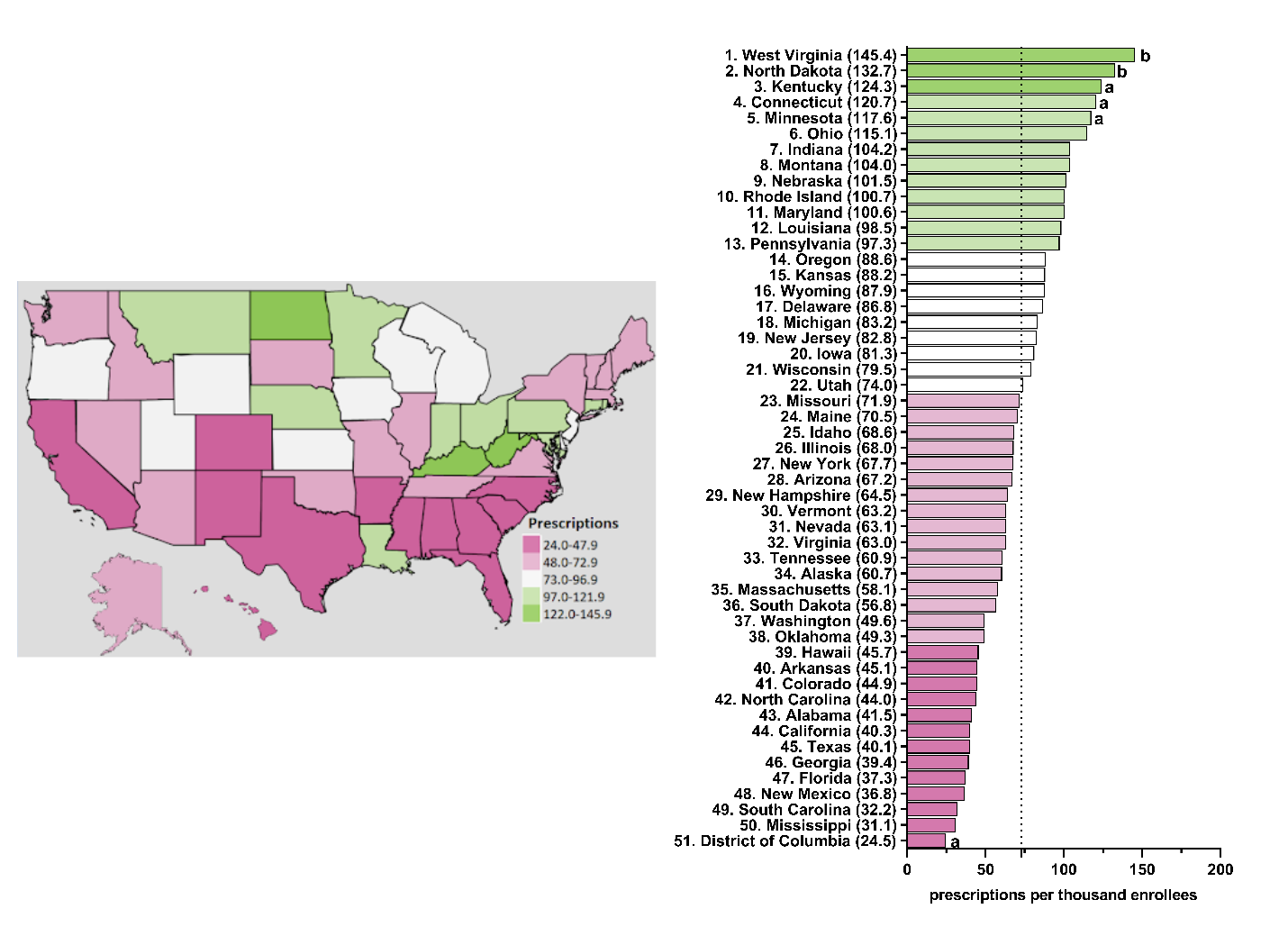

**Supplemental Figure 10.** Escitalopram prescriptions per thousand Medicaid enrollees heatmap (left) and population-corrected prescription rate per state (right) in 2019. ^a^ indicates ≥1.50 SD (32.5) from the mean (80.1). ^b^ indicates ≥1.96 SD from the mean.

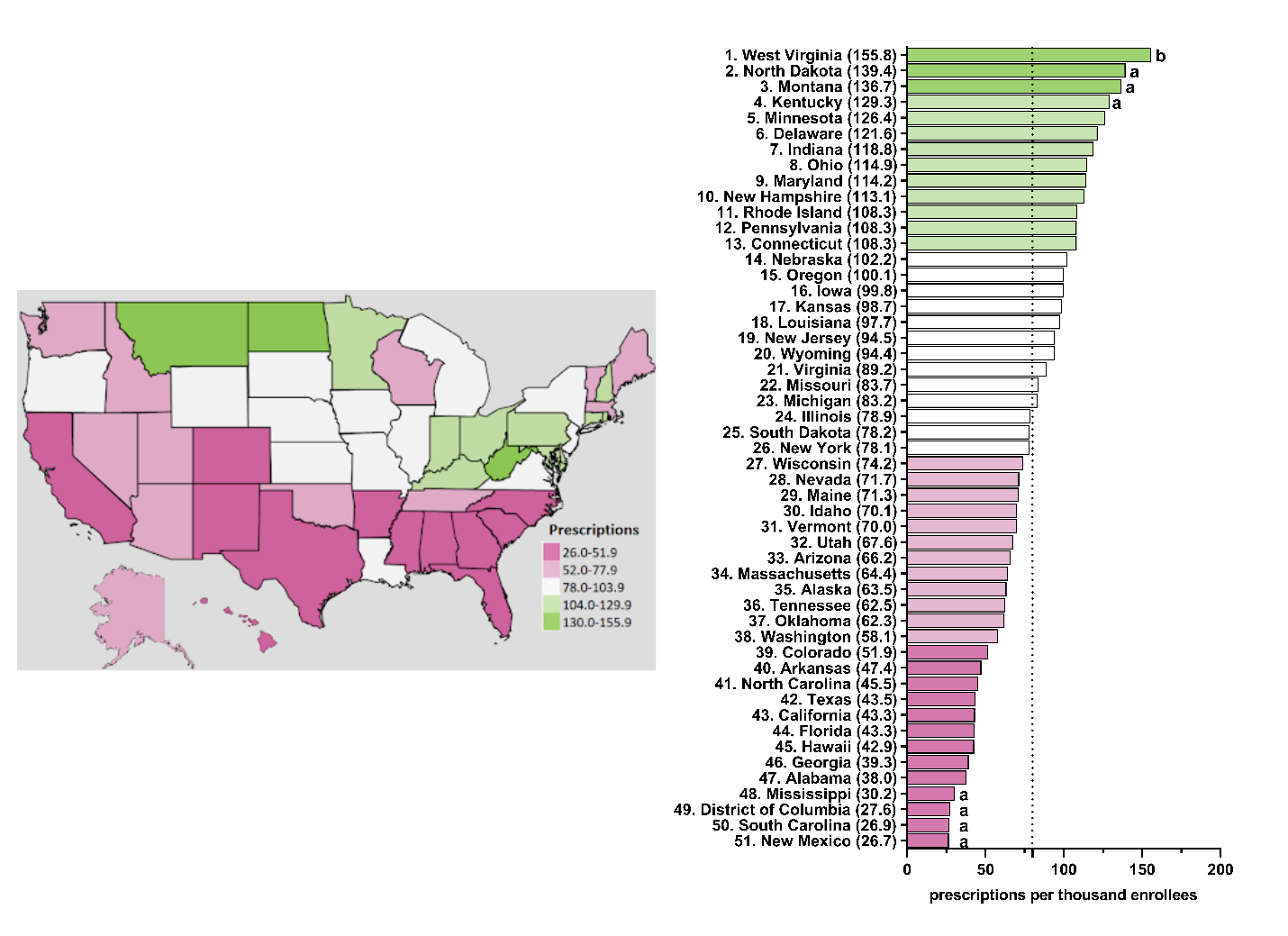

**Supplemental Figure 11.** Citalopram prescriptions per thousand Medicare enrollees heatmap (left) and population-corrected prescription rate per state (right) in 2015. ^a^ indicates ≥1.50 SD (81.0) from the mean (289.7). ^b^ indicates ≥1.96 SD from the mean.

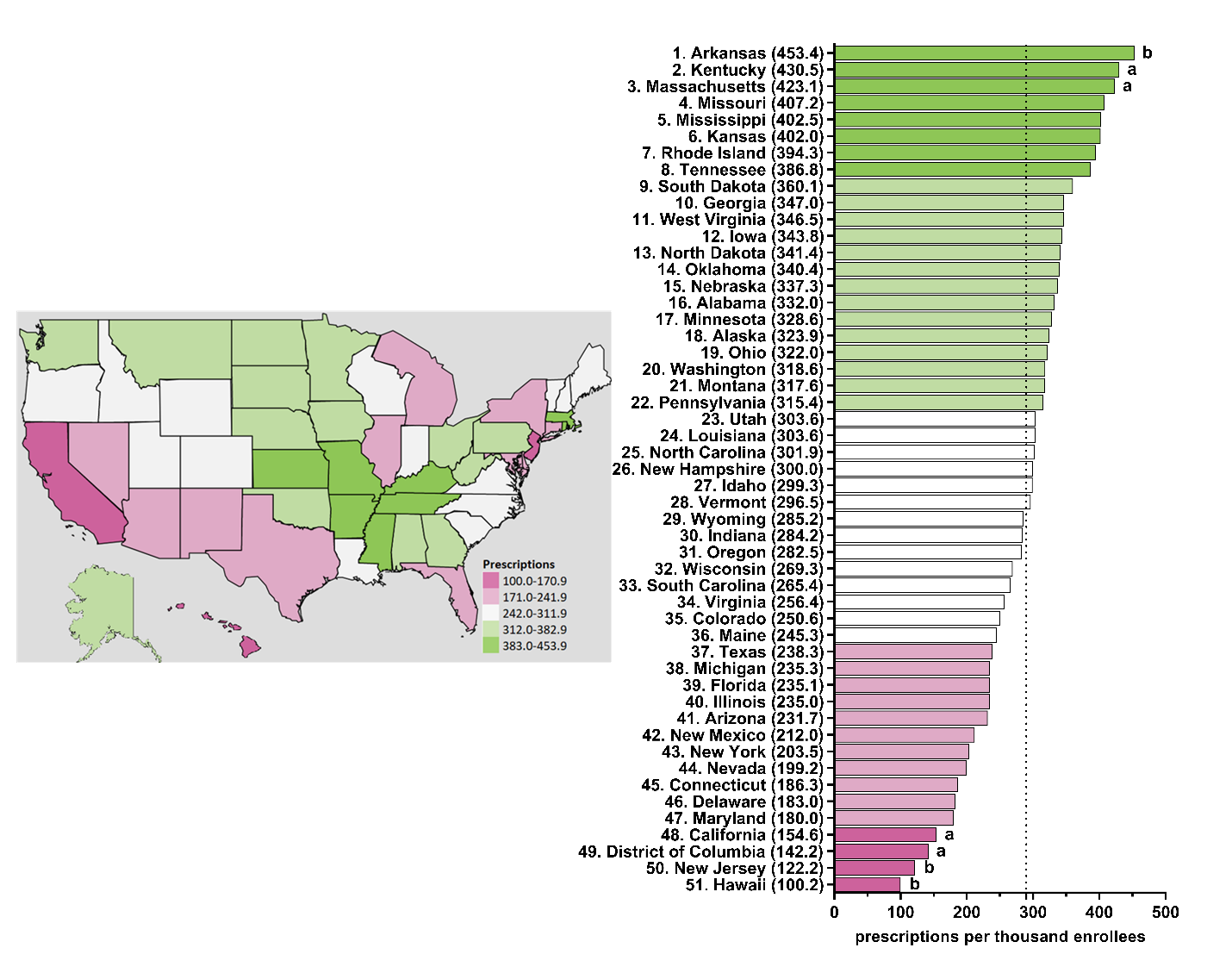

**Supplemental Figure 12.** Citalopram prescriptions per thousand Medicare enrollees heatmap (left) and population-corrected prescription rate per state (right) in 2016. ^a^ indicates ≥1.50 SD (78.7) from the mean (270.9). ^b^ indicates ≥1.96 SD from the mean.

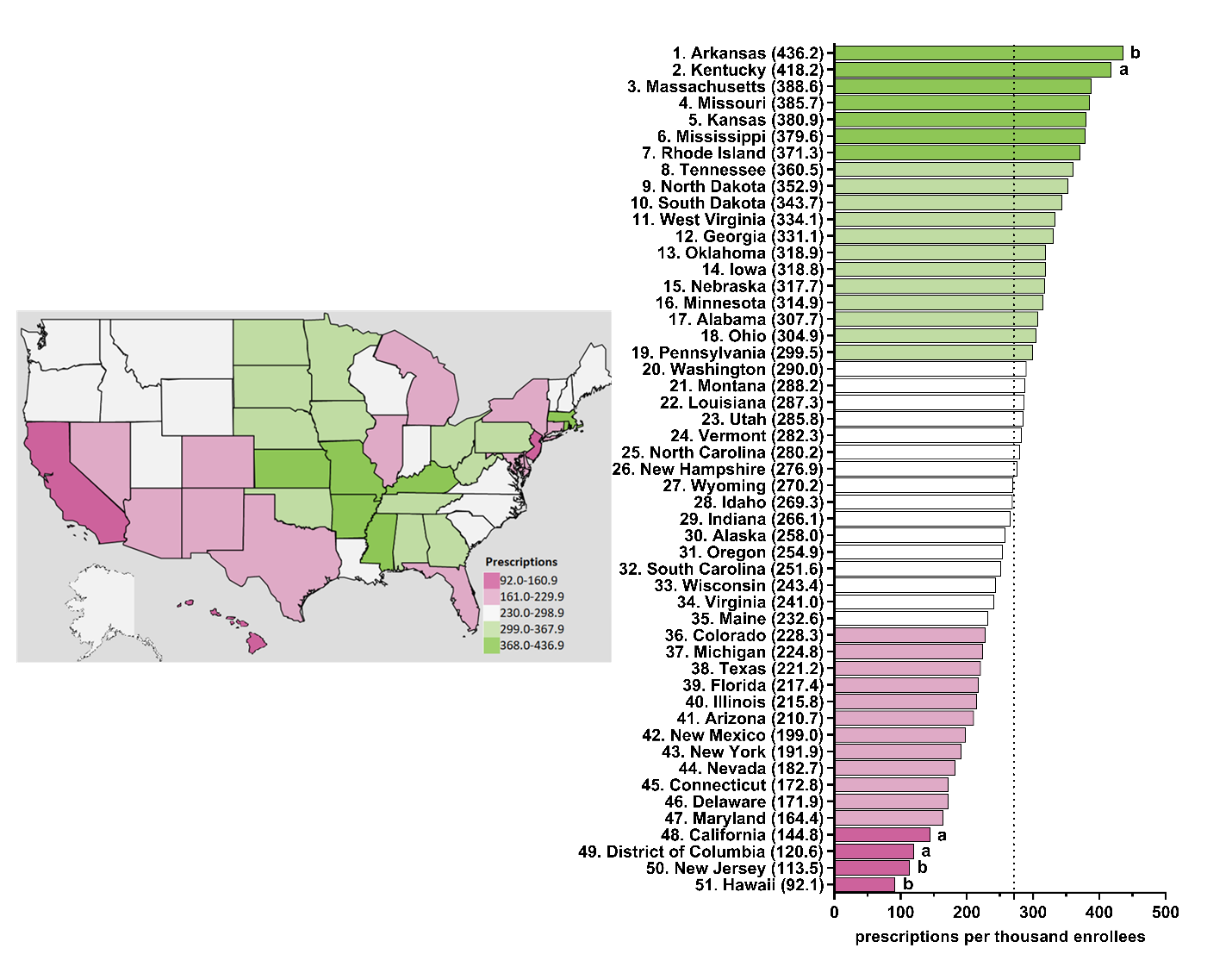

**Supplemental Figure 13.** Citalopram prescriptions per thousand Medicare enrollees heatmap (left) and population-corrected prescription rate per state (right) in 2017. ^a^ indicates ≥1.50 SD (72.7) from the mean (246.6). ^b^ indicates ≥1.96 SD from the mean.

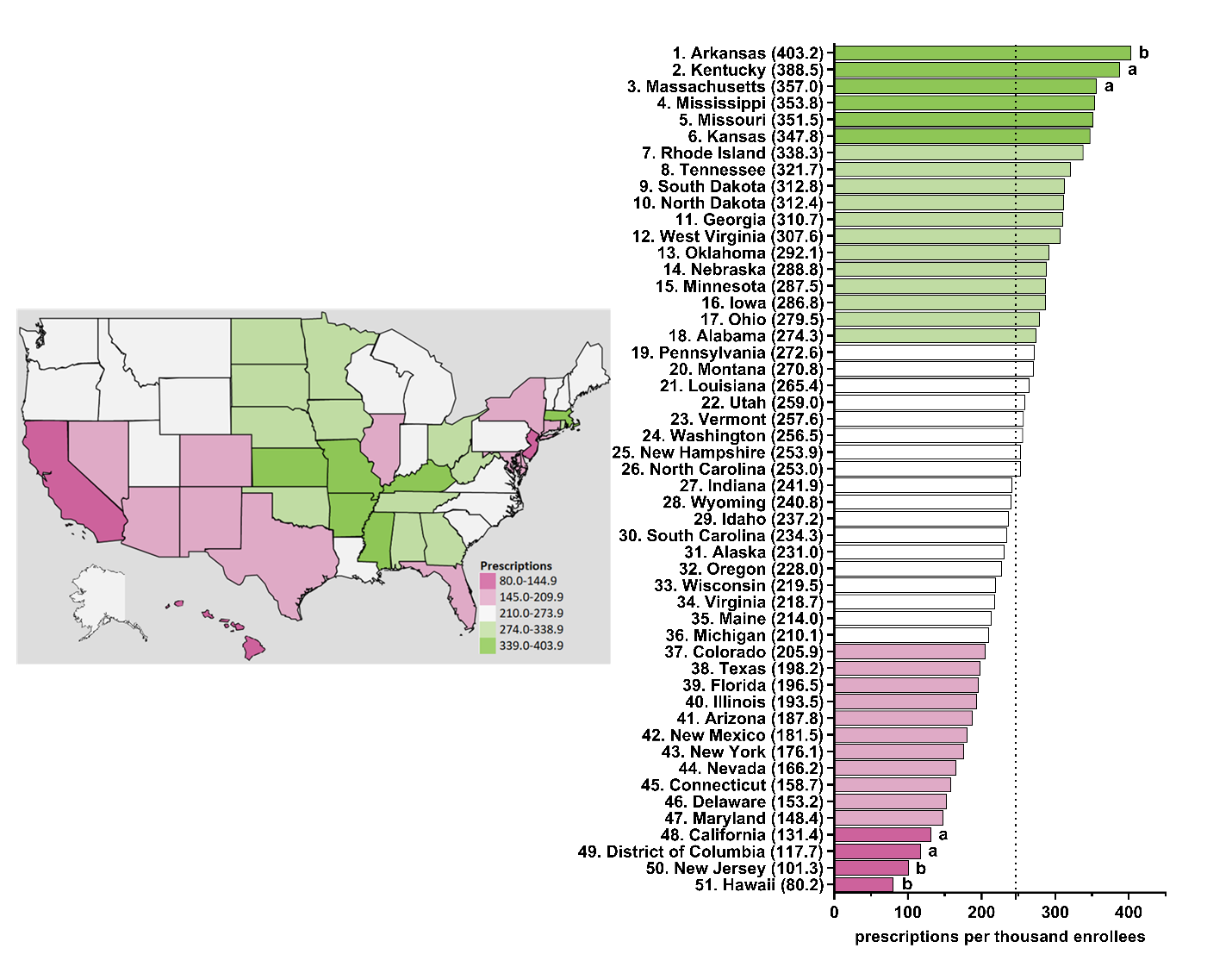

**Supplemental Figure 14.** Citalopram prescriptions per thousand Medicare enrollees heatmap (left) and population-corrected prescription rate per state (right) in 2018. ^a^ indicates ≥1.50 SD (65.4) from the mean (222.7). ^b^ indicates ≥1.96 SD from the mean.

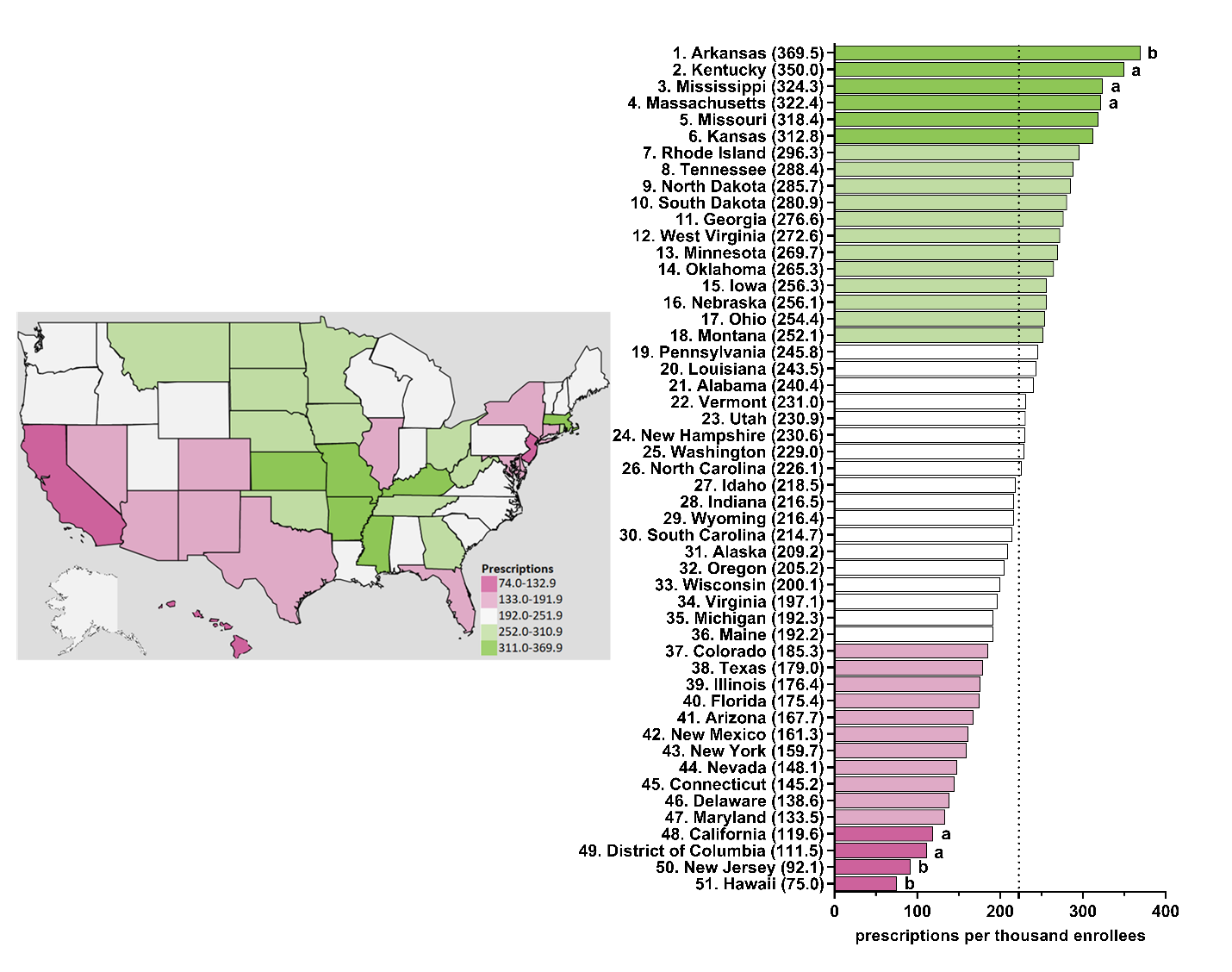

**Supplemental Figure 15.** Citalopram prescriptions per thousand Medicare enrollees heatmap (left) and population-corrected prescription rate per state (right) in 2019. ^a^ indicates ≥1.50 SD (57.6) from the mean (197.7). ^b^ indicates ≥1.96 SD from the mean.

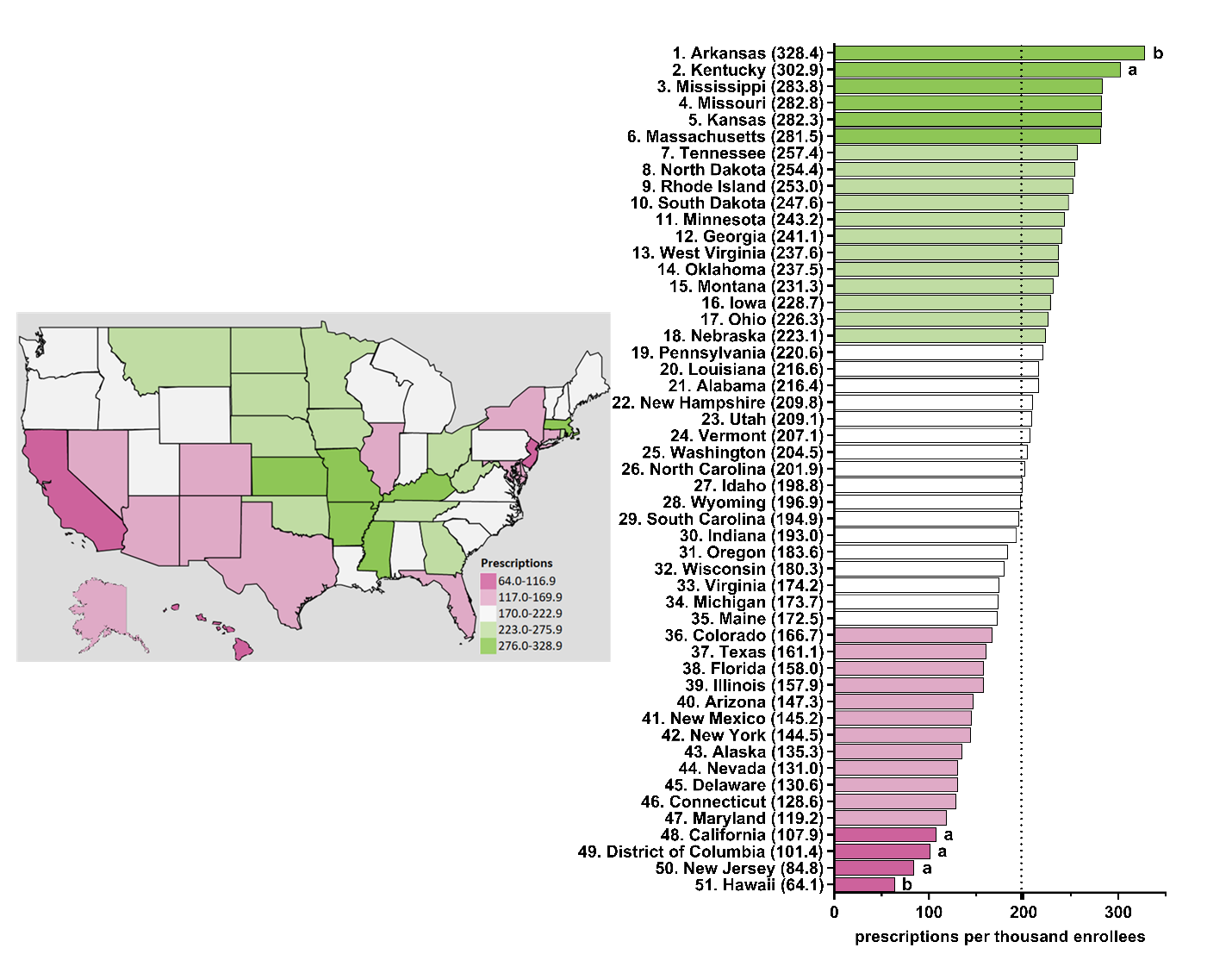

**Supplemental Figure 16.** Escitalopram prescriptions per thousand Medicare enrollees heatmap (left) and population-corrected prescription rate per state (right) in 2015. ^a^ indicates ≥1.50 SD (55.2) from the mean (210.2). ^b^ indicates ≥1.96 SD from the mean.

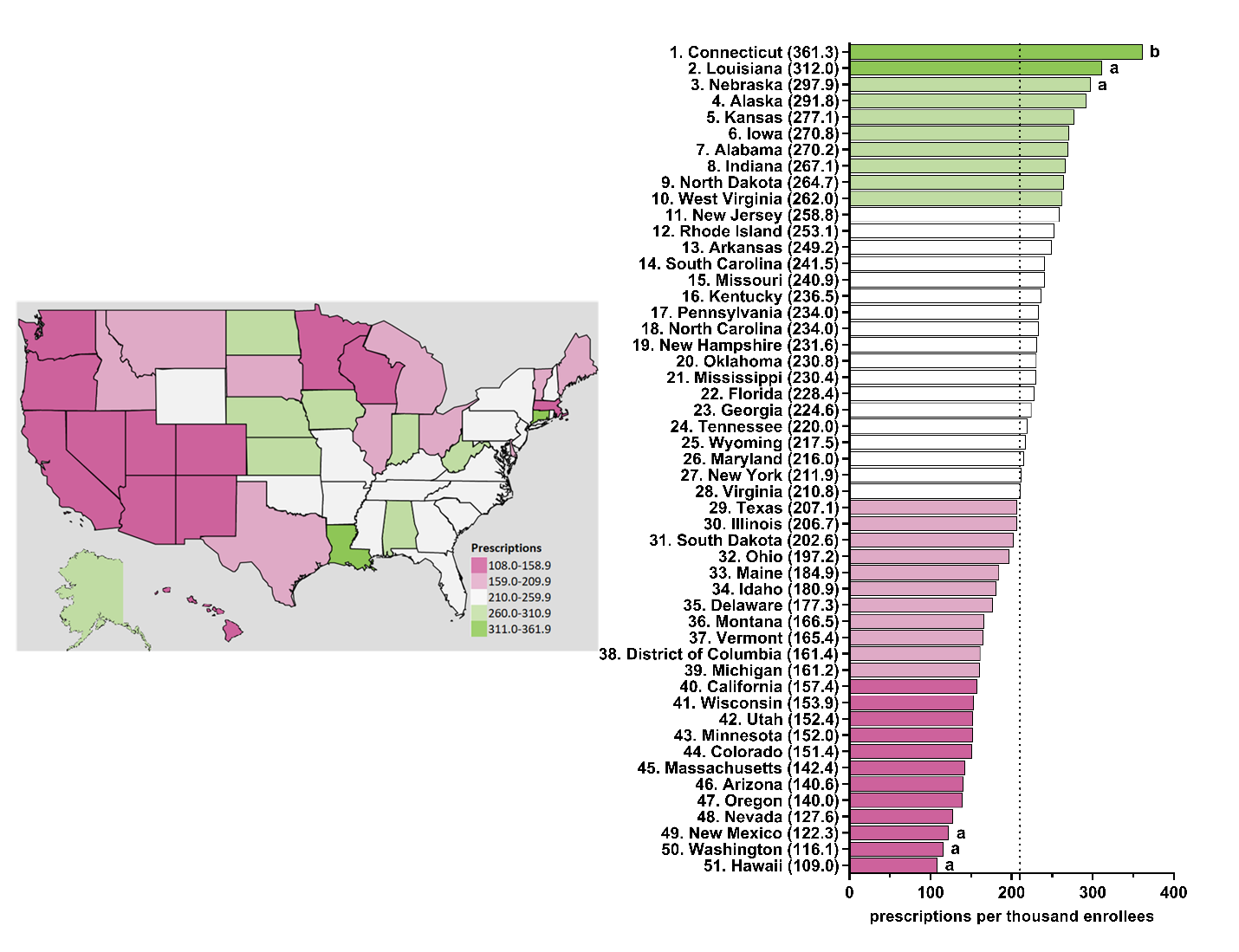

**Supplemental Figure 17.** Escitalopram prescriptions per thousand Medicare enrollees heatmap (left) and population-corrected prescription rate per state (right) in 2016. ^a^ indicates ≥1.50 SD (56.3) from the mean (218.9). ^b^ indicates ≥1.96 SD from the mean.

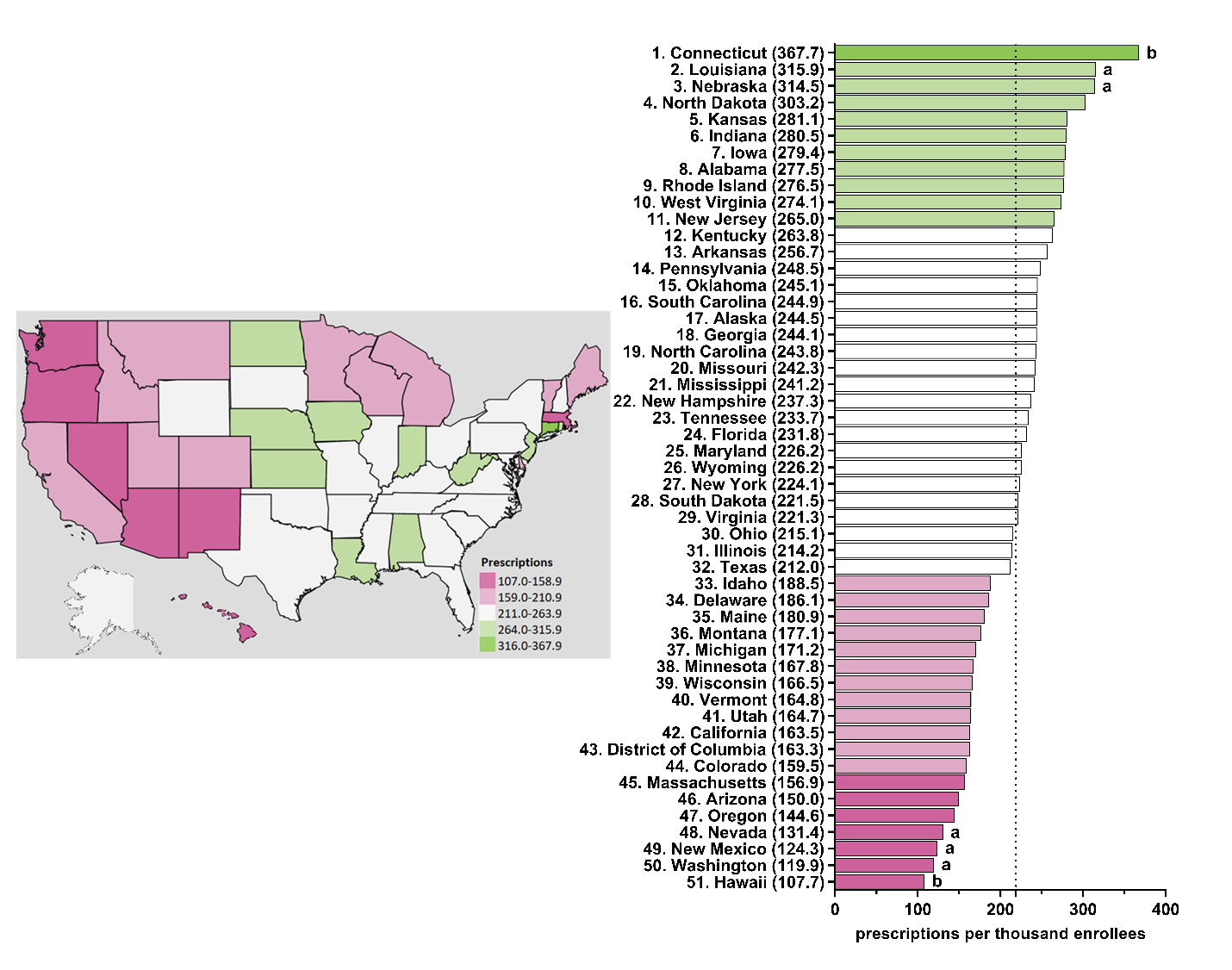

**Supplemental Figure 18.** Escitalopram prescriptions per thousand Medicare enrollees heatmap (left) and population-corrected prescription rate per state (right) in 2017. ^a^ indicates ≥1.50 SD (56.2) from the mean (222.5). ^b^ indicates ≥1.96 SD from the mean.

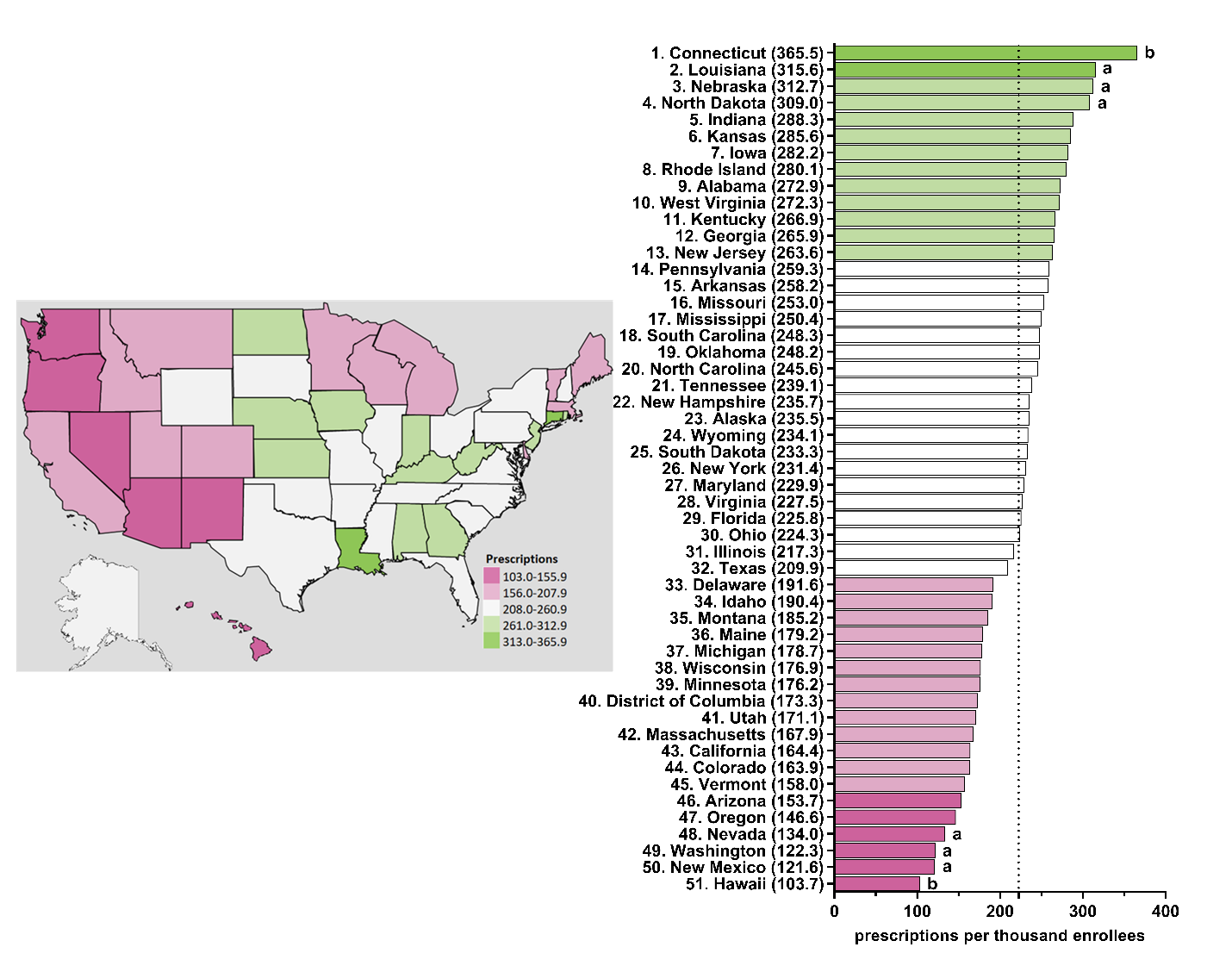

**Supplemental Figure 19.** Escitalopram prescriptions per thousand Medicare enrollees heatmap (left) and population-corrected prescription rate per state (right) in 2018. ^a^ indicates ≥1.50 SD (54.2) from the mean (222.1). ^b^ indicates ≥1.96 SD from the mean.

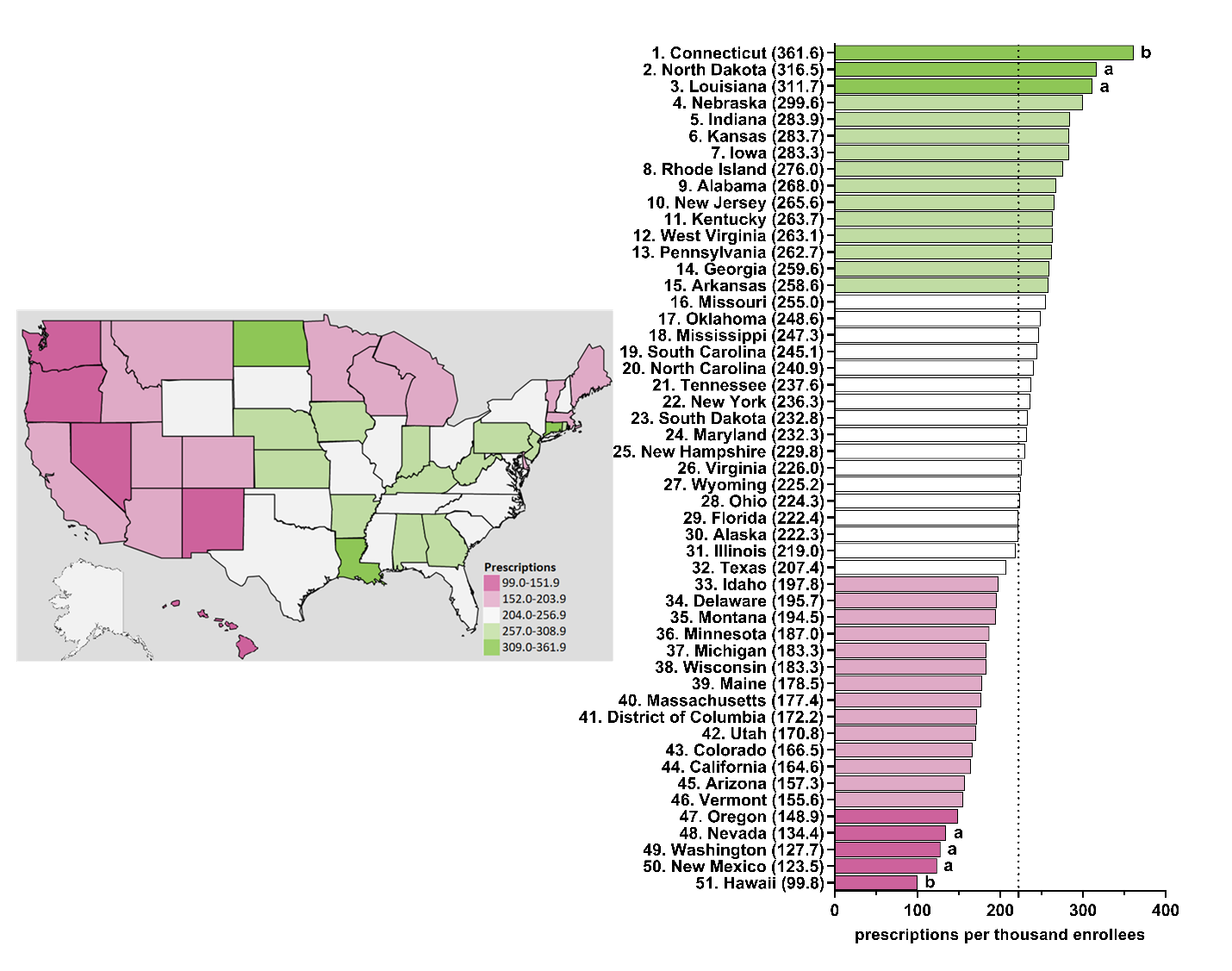

**Supplemental Figure 20.** Escitalopram prescriptions per thousand Medicare enrollees heatmap (left) and population-corrected prescription rate per state (right) in 2019. ^a^ indicates ≥1.50 SD (52.0) from the mean (218.9). ^b^ indicates ≥1.96 SD from the mean.

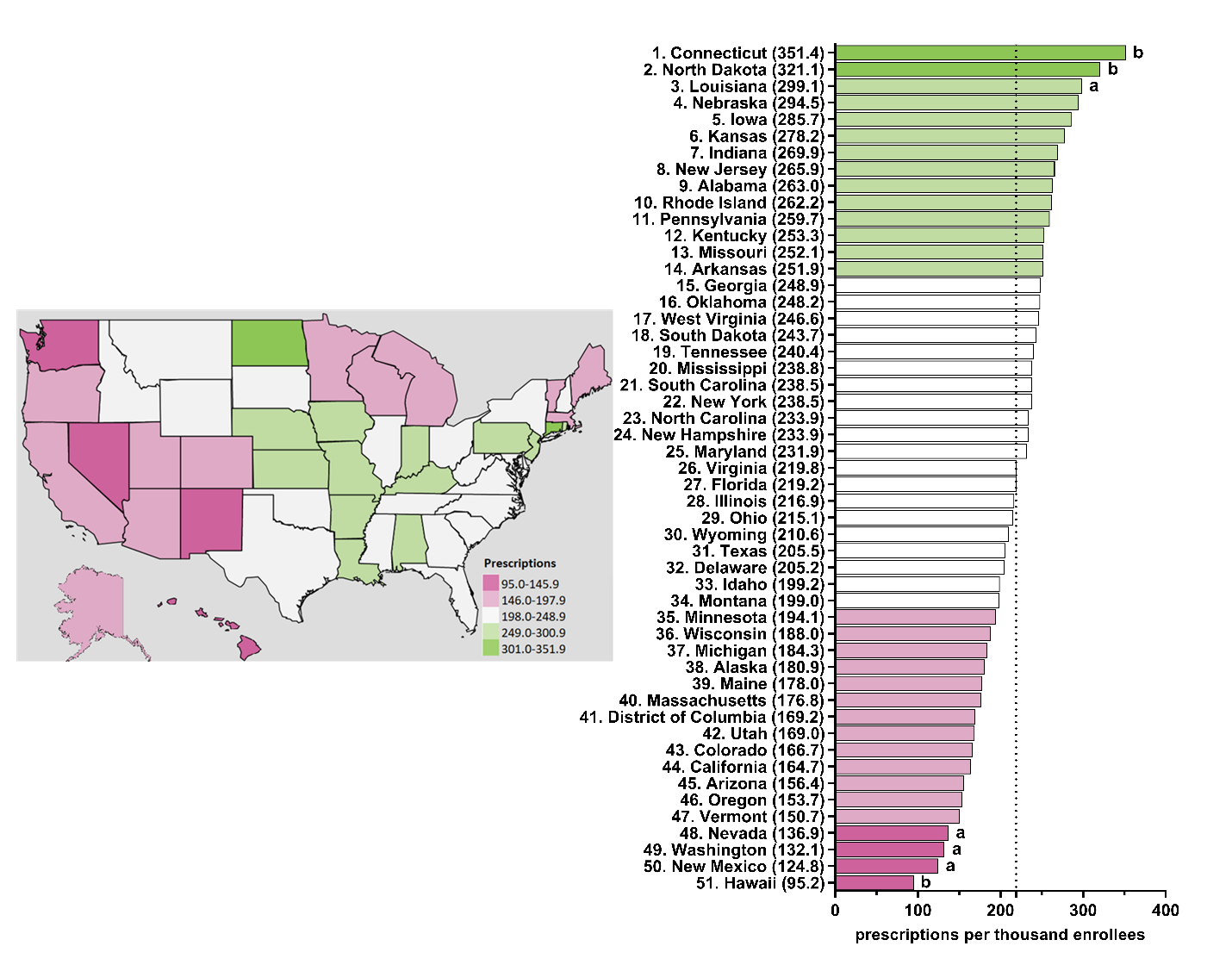

**Supplemental Figure 21**. Matrix of Pearson’s correlation coefficients between population-corrected number of prescriptions of citalopram and escitalopram within the Medicaid and Medicare systems for 2015 (*N*=51: 50 states and D.C.). * indicates *p*≤0.05, ** indicates *p*≤0.01, and *** *p*≤0.001.

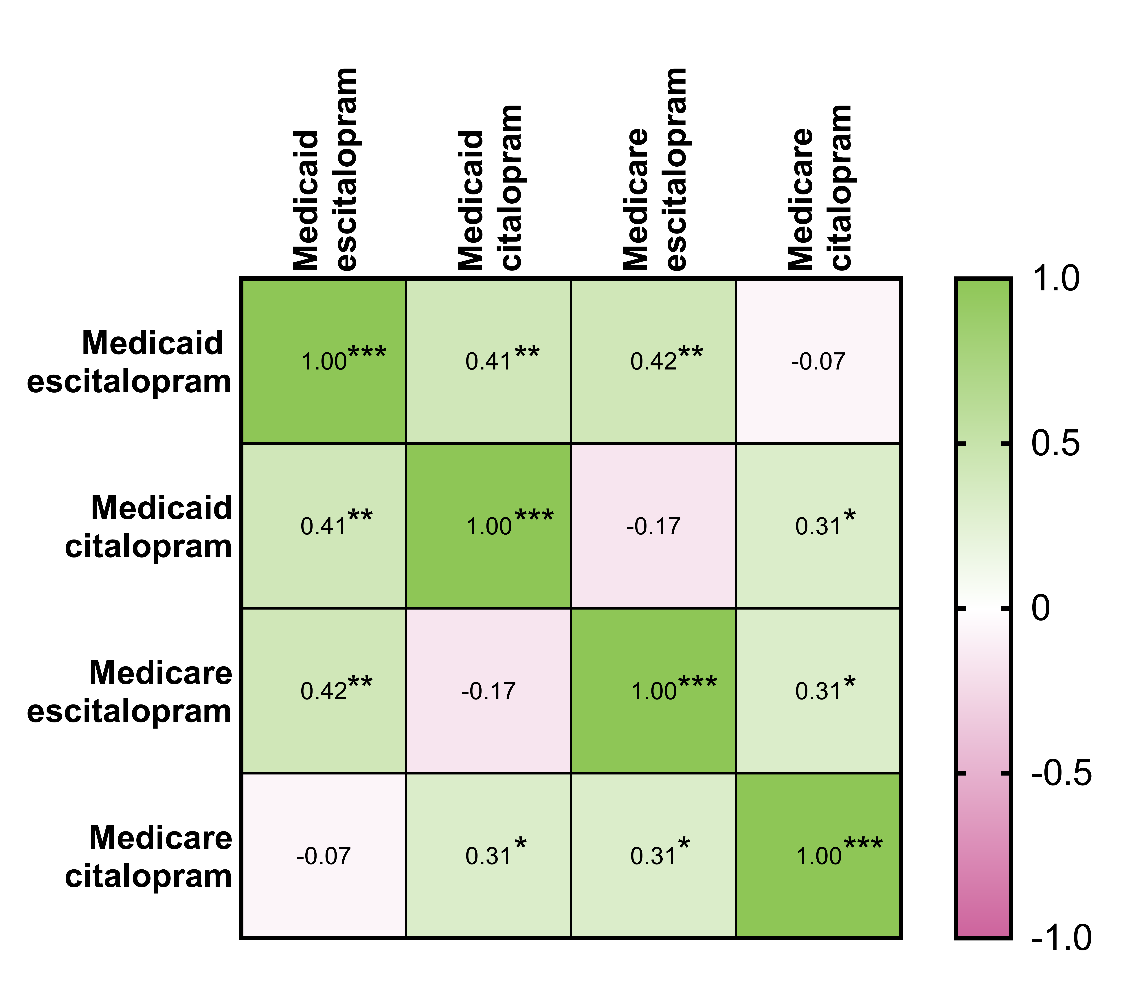

**Supplemental Figure 22**. Matrix of Pearson’s correlation coefficients between population-corrected number of prescriptions of citalopram and escitalopram within the Medicaid and Medicare systems for 2016 (*N*=51: 50 states and D.C.). * indicates *p*≤0.05, ** indicates *p*≤0.01, and *** *p*≤0.001.

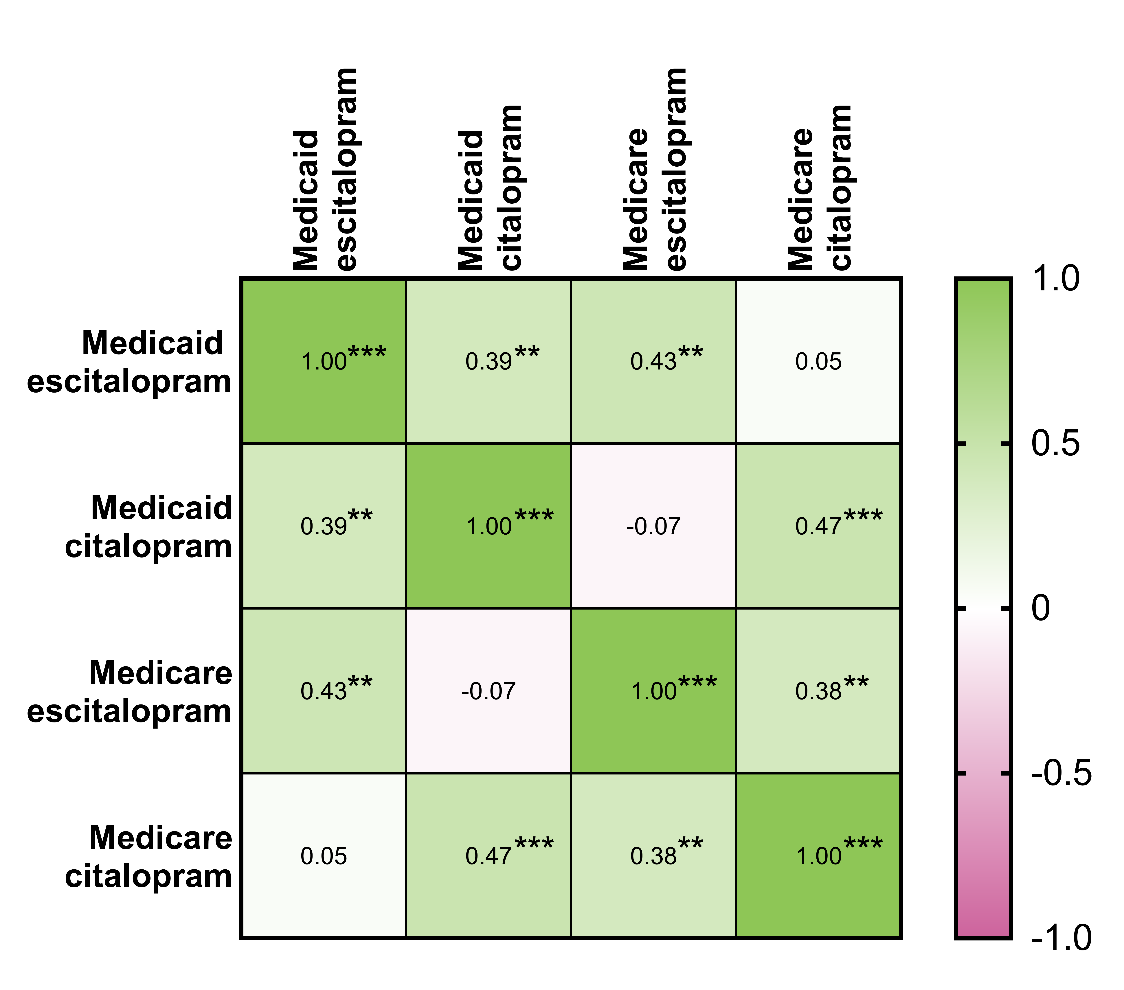

**Supplemental Figure 23**. Matrix of Pearson’s correlation coefficients between population-corrected number of prescriptions of citalopram and escitalopram within the Medicaid and Medicare systems for 2017 (*N*=51: 50 states and D.C.). * indicates *p*≤0.05, ** indicates *p*≤0.01, and *** *p*≤0.001.

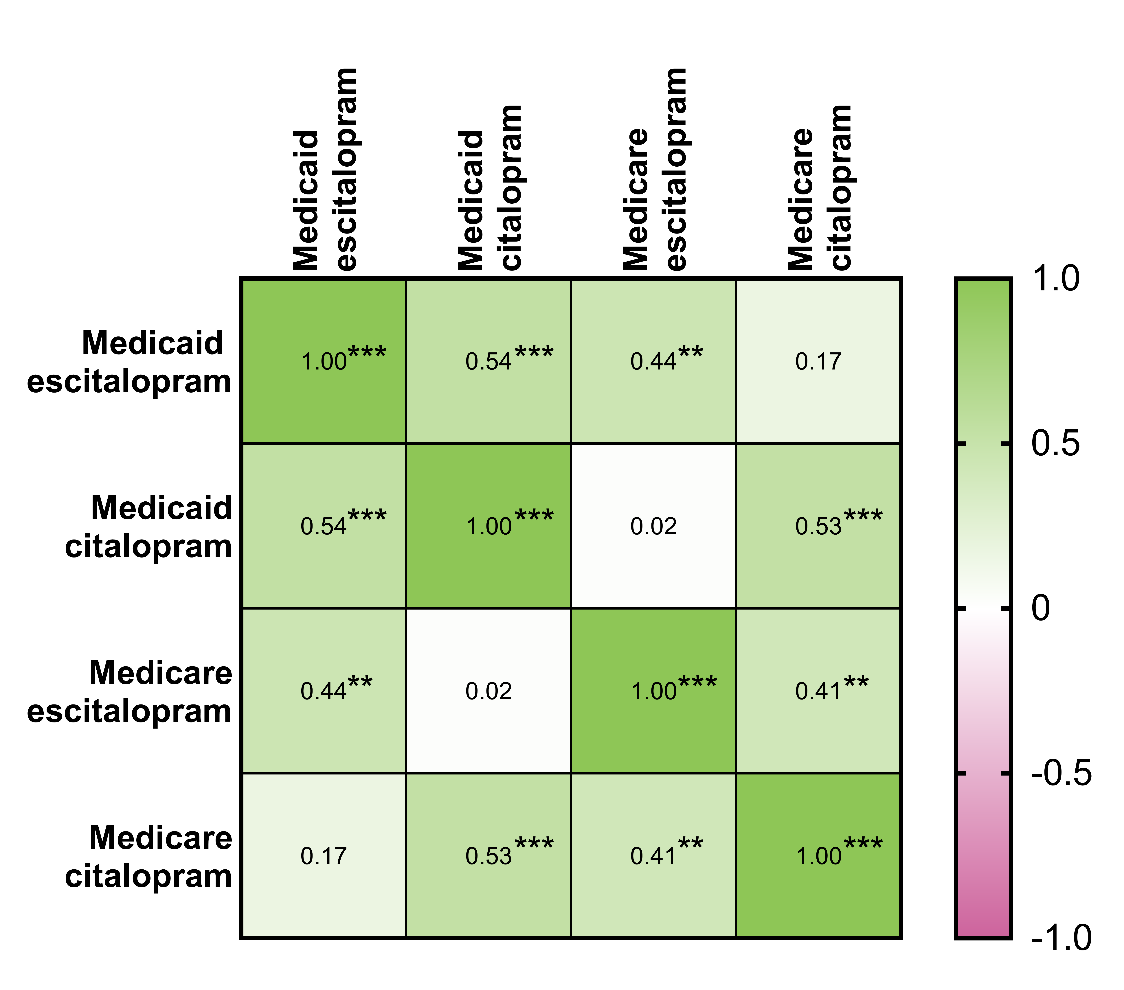

**Supplemental Figure 24**. Matrix of Pearson’s correlation coefficients between population-corrected number of prescriptions of citalopram and escitalopram within the Medicaid and Medicare systems for 2018 (*N*=51: 50 states and D.C.). * indicates *p*≤0.05, ** indicates *p*≤0.01, and *** *p*≤0.001.

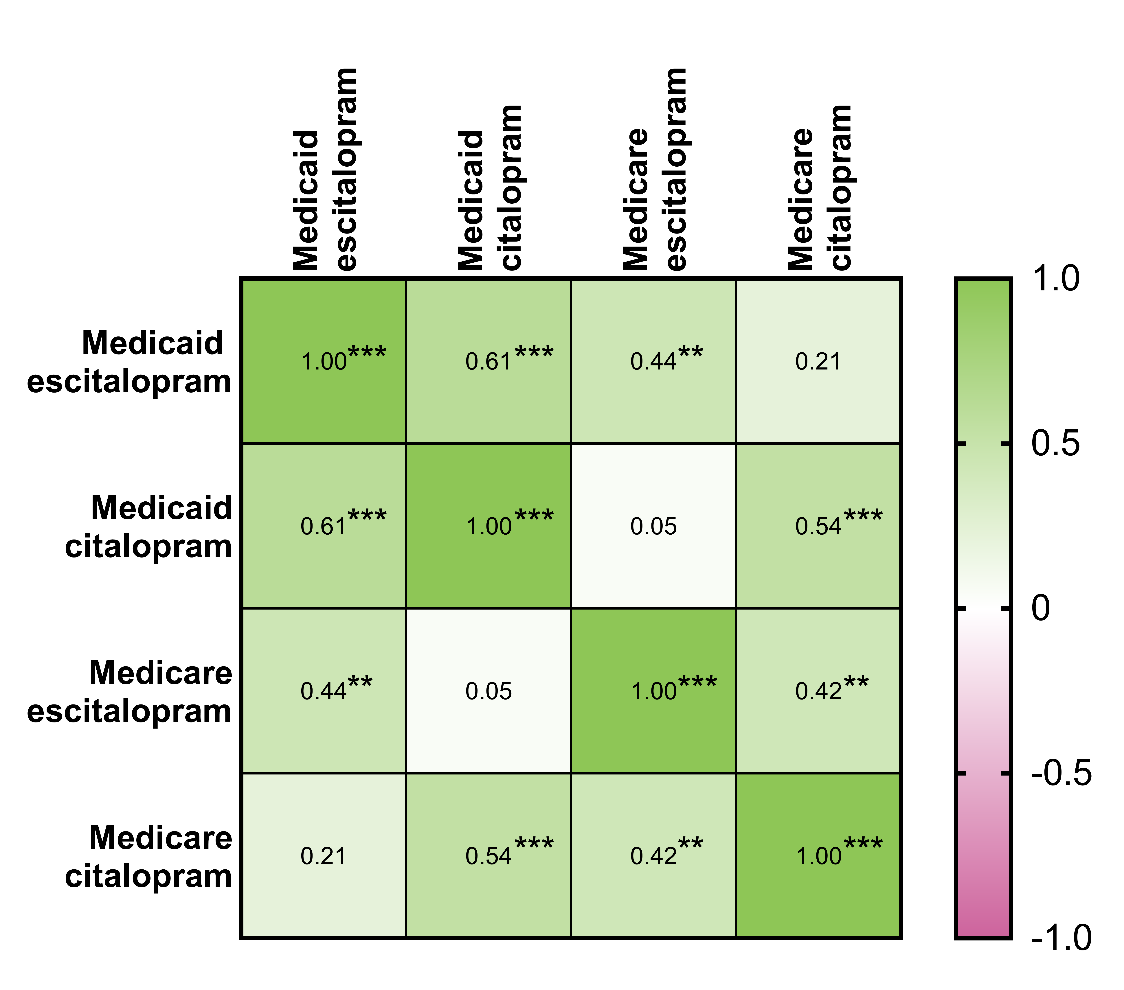

**Supplemental Figure 25**. Matrix of Pearson’s correlation coefficients between population-corrected number of prescriptions of citalopram and escitalopram within the Medicaid and Medicare systems for 2019 (*N*=51: 50 states and D.C.). * indicates *p*≤0.05, ** indicates *p*≤0.01, and *** *p*≤0.001.

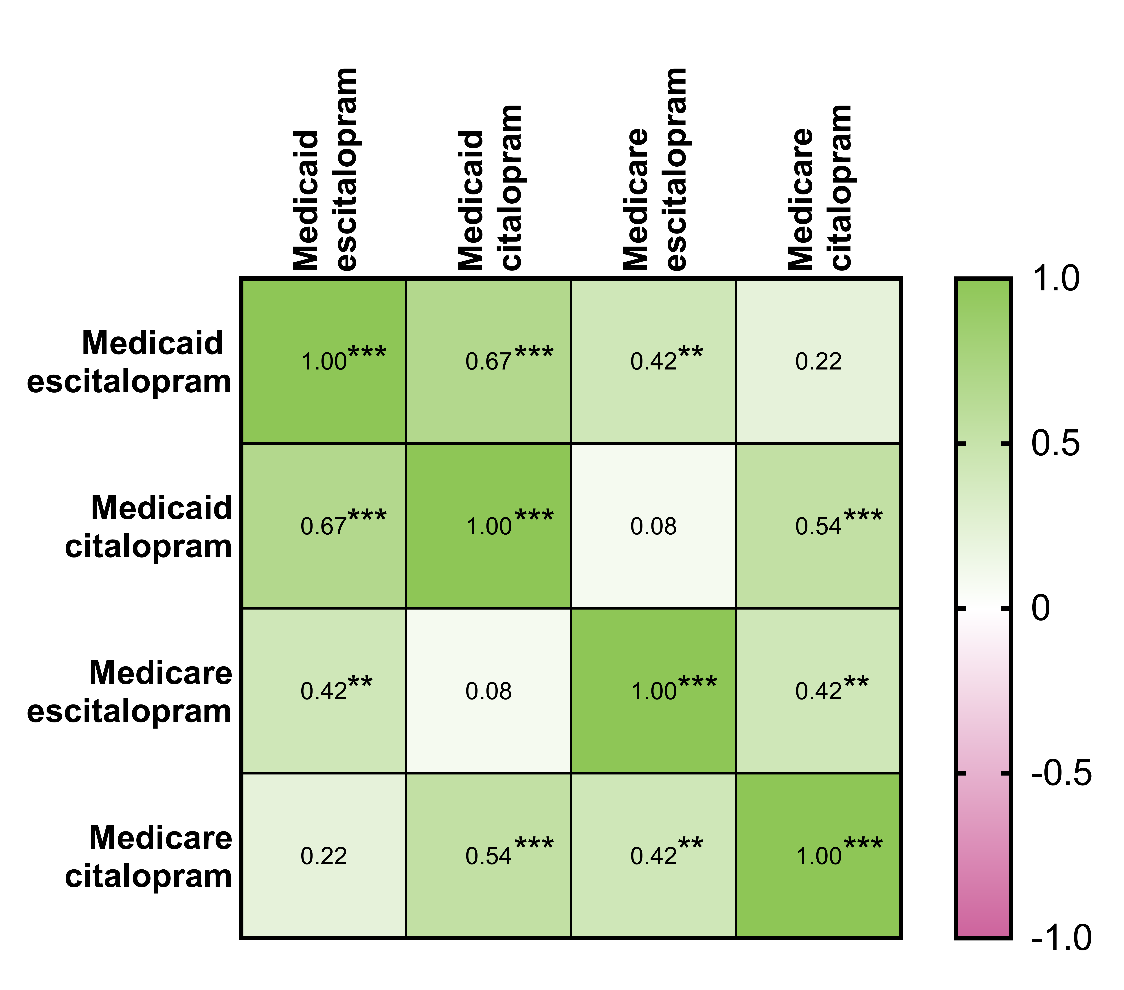
